# Risk Factors for Antimicrobial Resistance in Cancer Patients and Cancer Survivors: An Electronic Health Record Study

**DOI:** 10.64898/2026.04.17.26351097

**Authors:** Feifei Hu, Jia Wei, Berit Muller-Pebody, Russell Hope, Colin Brown, Helena Carreira, Alicia Demirjian, A. Sarah Walker, David W. Eyre

## Abstract

**Objectives:** To identifiy risk factors for antimicrobial resistance (AMR) in seven pathogen-antimicrobial combinations in patients with cancer and cancer survivors.

**Methods:** Using data from patients with recent or past cancer diagnostic codes in Oxfordshire, UK, we examined associations between 22 potential risk-factors and AMR in blood culture isolates, collected between 1-April-2015 and 31-March-2025.

**Results:** Among 5,975 bacteraemias in 4,365 adults, we analysed 3,141 (52.6%) due to Enterobacterales and 620 (10.4%) due to *Enterococcus faecalis/faecium* in 2,752 patients. Fourteen risk-factors for antimicrobial-resistant bacteraemia were identified, varying across pathogen-antimicrobial combinations. Compared with no previous antimicrobial susceptibility test result, prior resistance to the same antibiotic in any culture in the last year was strongly associated with AMR across all pathogen-antimicrobial combinations (all p≤0.001). Prior antibiotic exposure and younger age were also positively associated with AMR in four and five combinations, respectively. Cancer type showed modest effects; lymphoid/haematopoietic malignancies were associated with higher odds (vs colorectal cancer) of trimethoprim-sulfamethoxazole-resistant Enterobacterales (aOR=2.07 95%CI 1.40-3.06) and vancomycin-resistant *Enterococcus* bacteraemia (aOR=6.68, 1.21-36.91).

**Conclusions:** Previous resistance was the greatest risk factor for bacteraemia with AMR in cancer patients and survivors, with prior antibiotic exposure and age also contributing. Lymphoid/haematopoietic malignancies increased risk of resistance to specific antimicrobials. Keywords: antimicrobial resistance, bacteraemia, cancer, risk factors

## Introduction

Antimicrobial resistance (AMR) is an increasing healthcare challenge and has been recognised as a global public health threat by the World Health Organization^1^. As AMR decreases the effectiveness of many antimicrobials, routine infections become more difficult to manage, increasing morbidity and pressure on health systems^2^. In England, around 400 new antibiotic resistant infections were reported each week in 2024^3^. People undergoing cancer treatment are especially vulnerable to infections with potentially longer lasting effects in cancer survivors as well. Both the underlying malignancy and the effects of chemotherapy and other treatments affect immune function^4^. Infections remain a major contributor to mortality in patients with cancer, with some studies suggesting over half of cancer-related deaths involve infections^4^. Additionally, the proportion of infections with AMR in patients with cancer is increasing^5, 6^, e.g., leading to increases in mortality among patients receiving chemotherapy for haematological malignancies^7^. In England, rates of fatal bacteraemia caused by resistant organisms have risen since 2023, despite reductions in National Health Service (NHS) antimicrobial prescribing^3^; in 2024, 85.1% of resistant bacteraemias were caused by Enterobacterales^8^.

Given this context, identifying factors increasing AMR risk in patients with cancer and in cancer survivors is essential for improving outcomes from infection, and supporting prevention strategies and antimicrobial stewardship. Previous work has highlighted several AMR risk factors including prior antibiotic exposure, previous hospital stays, urinary catheterisation^9^, urinary-tract infections, comorbidities^10^, and previous resistant bacterial cultures^11^. However, most studies have focused on single pathogens or limited antimicrobial classes, and few have compared multiple risk factors across different ‘bug-drug’ groups. In addition, earlier research has often considered only prior resistance to the same antimicrobial or in the same type of culture as potential risk factors, overlooking potential epidemiological and mechanistic associations with resistance to other drug classes or resistance detected in different sample types.

To address these gaps, and to generate data for cancer survivors as well as patients with cancer, we used electronic health record data to examine a broad set of demographical, clinical and microbiological factors for associations with AMR across seven key pathogen-antimicrobial combinations, including evaluating prior resistance to both target and non-target antimicrobials in any sample type (including both blood and non-blood cultures), alongside previous susceptibility. The aim was to obtain evidence to support more targeted AMR interventions and enhance protection for high-risk cancer populations.

## Methods

We used data from the Infections in Oxfordshire Research Database (IORD), which includes de-identified medical records from four hospitals comprising the Oxford University Hospitals (OUH), results from samples sent from the community and hospital for testing at the OUH laboratories, and hospital prescribing and details of all cancer chemotherapy and radiotherapy.

These hospitals serve a population of approximately 755,000 individuals. IORD has approvals from the National Research Ethics Service South Central Oxford C Research Ethics Committee (24/SC/0241), the Health Research Authority, and the Confidentiality Advisory Group (19/CAG/0144) as a research database not requiring individual patient consent.

We studied all patients with a primary or secondary ICD-10 diagnostic code consistent with cancer (including C00-C97) between 1 April 2000 and 31 March 2025 who were aged ≥16 years at the first such code, i.e., studying both current cancer patients and cancer survivors.

The primary outcome was antimicrobial resistance (AMR) vs susceptibility among positive blood cultures (denoted ‘bacteraemia’) in these patients between 1 April 2015 and 31 March 2025. AMR was identified based on routine diagnostic testing performed by OUH microbiology laboratory using European Committee on Antimicrobial Susceptibility Testing^12^ (EUCAST) standards. Bacteraemias were deduplicated by considering repeat positive blood cultures of the same pathogen within 14 days of an initial positive blood culture as the same infection episode. Where multiple antimicrobial susceptibility test (AST) results for a given pathogen-antimicrobial combination were available within the 14 day window the episode was considered resistant if any result for the antibiotic was resistant, susceptible if all were susceptible, otherwise unknown if any result for the antibiotic was unknown. Analyses focused on the seven pathogen-antimicrobial combinations with the largest sample sizes, namely resistance to third-generation cephalosporins, co-amoxiclav, fluoroquinolones, trimethoprim/sulfamethoxazole, gentamicin, and piperacillin-tazobactam in Enterobacterales, as well as resistance to vancomycin in *Enterococcus faecalis/faecium* (**Table 1**, **Figure 1**).

**Figure 1:**
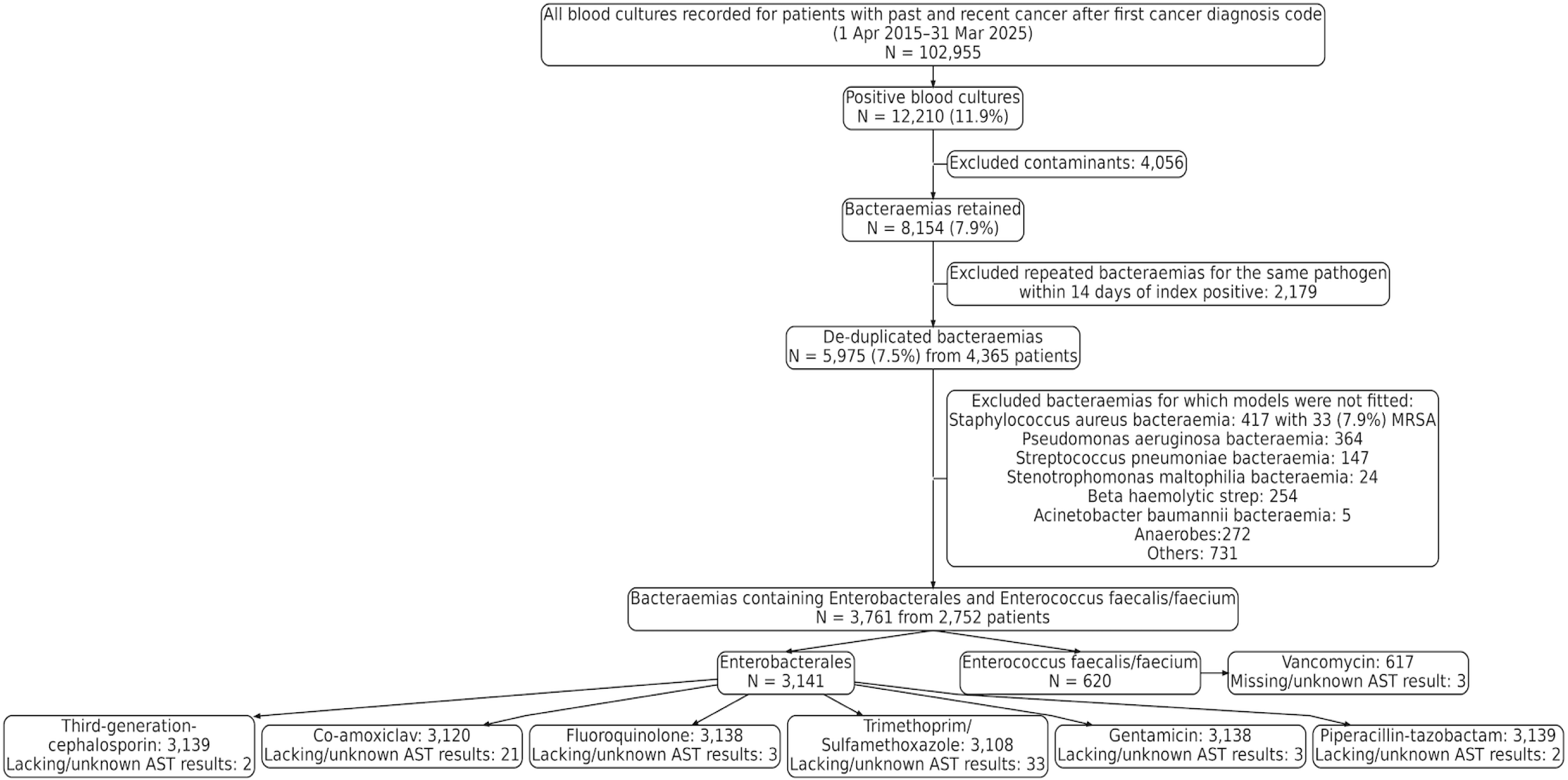
Study flow chart. There were only 40/3138 (1.3%) Enterobacterales infections resistant to any carbapenem which were not included in analyses. A list of contaminants is provided in the **Supplementary Methods**.

**Table 1:**
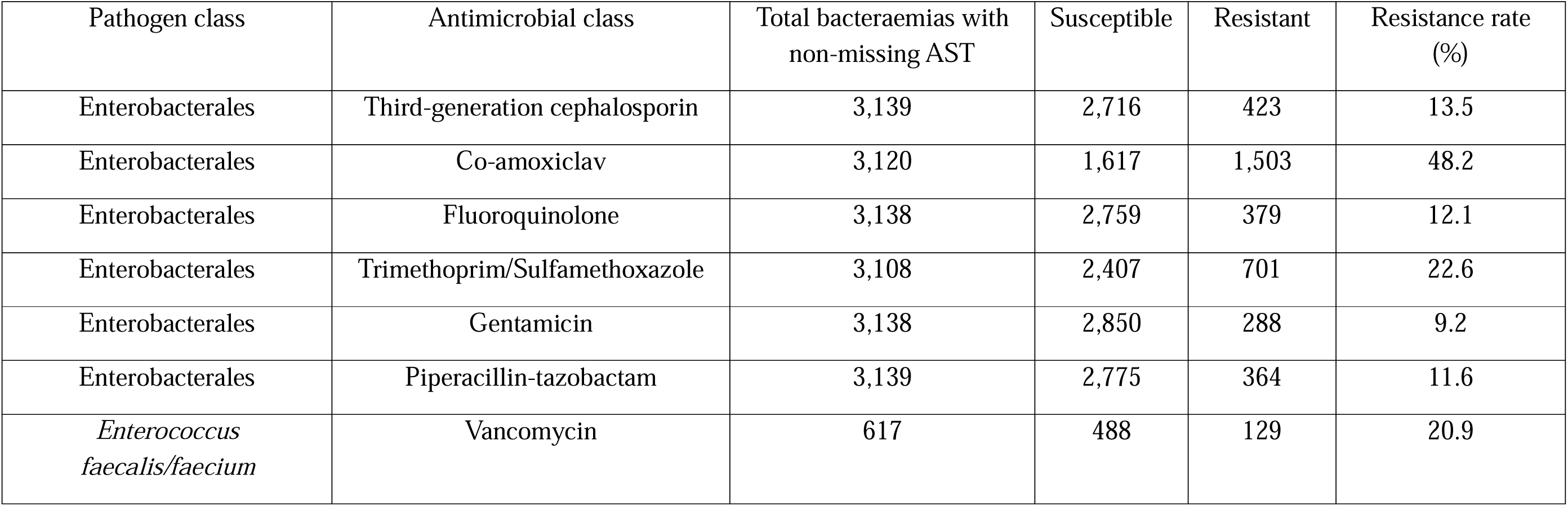
Resistance summary in included pathogen-antimicrobial combinations. AST=antimicrobial susceptibility.

| Pathogen class | Antimicrobial class | Total bacteraemias with non-missing AST | Susceptible | Resistant | Resistance rate (%) |
| --- | --- | --- | --- | --- | --- |
| Enterobacterales | Third-generation cephalosporin | 3,139 | 2,716 | 423 | 13.5 |
| Enterobacterales | Co-amoxiclav | 3,120 | 1,617 | 1,503 | 48.2 |
| Enterobacterales | Fluoroquinolone | 3,138 | 2,759 | 379 | 12.1 |
| Enterobacterales | Trimethoprim/Sulfamethoxazole | 3,108 | 2,407 | 701 | 22.6 |
| Enterobacterales | Gentamicin | 3,138 | 2,850 | 288 | 9.2 |
| Enterobacterales | Piperacillin-tazobactam | 3,139 | 2,775 | 364 | 11.6 |
| *Enterococcus faecalis/faecium* | Vancomycin | 617 | 488 | 129 | 20.9 |

Potential risk factors were selected from previous studies of AMR in any population^13–16^. These included, at the bacteraemia index date, patient demographics and comorbidities (age, sex, ethnicity, deprivation percentiles^17^, Charlson comorbidity score^18, 19^, **Supplementary Methods**); cancer-related factors, including type (breast, prostate, colorectal, upper gastrointestinal and hepatobiliary, melanoma, non-melanoma skin, lung, lymphoid/haematopoietic, and other cancers; based on the first ICD-10 code where patients were diagnosed with multiple cancers), time since first cancer diagnostic code, cancer treatment in the past year based on presence of any radiotherapy/chemotherapy treatment record or procedure codes (OPCS-4 codes: X65, X67-69, X72-73), and number of neutrophil tests taken in the past year; clinical history within the last year, including frailty score based on ICD-10 codes^20^, counts of positive urine cultures with the specific pathogen of interest, counts of prior urine and blood samples sent for culture, prior positive catheter specimen urine (CSU) cultures, total days in hospital and in an intensive care unit (ICU); healthcare-associated factors, including if the bacteraemia was defined as hospital-onset, i.e., >48h after admission and before discharge, otherwise community-onset, calendar time since 1 April 2015, prior antibiotic resistance and susceptibility status for the specific pathogen groupings in **Table 1** in both blood and non-blood (i.e., all other) cultures in the last year; and prior antibiotic exposures in the last year (including antibiotics received in hospital and provided at discharge, but not primary care prescriptions), including cumulative days on any and specific antibiotics, and days since last antibiotic use. Since very few cultures were resistant to the target antibiotic but susceptible to all other antibiotics and as results from blood and non-blood cultures were highly congruent, we considered 4 previous ‘resistance’ categories based on the corresponding pathogen in any cultures in the last year: no previous AST result (i.e., no previous positive culture with the pathogen), any isolate with resistance to the target antimicrobial, any isolate with resistance but only to non-target antimicrobials, fully susceptible to all tested antimicrobials. In total, 22 variables were included (**Table S1**) (detailed definitions in **Supplementary Methods)**.

### Statistical analysis

For each pathogen-antimicrobial combination, we used logistic regression to estimate associations between resistant vs susceptible bacteraemia and all potential risk factors (listed in **Table S1**) using patient-clustered robust standard errors^21^. We used complete cases as data were complete for nearly all variables, with ∼0.5% observations missing deprivation scores. No patients with melanoma had vancomycin-resistant *Enterococcus* bacteraemia, so patients with this cancer were removed from this pathogen-antimicrobial model. Collinearity was evaluated using pairwise correlation, and one of each pair of potentially collinear variables (Spearman correlation >0.8^22^) were removed prior to model fitting based on clinical relevance. Non-linearity in continuous factors was incorporated where Bayesian information criterion (BIC) supported a superior fit with natural cubic splines (**Supplementary Methods**). Model selection then used backward elimination with exit p=0.05 for main effects and 0.01 for non-linear terms (always retaining linear terms until they were the only term being considered), forcing cancer type into all models. After constructing this final main effects model, we then evaluated pairwise interactions using forward selection based on Bonferroni-adjusted p≤0.05/number of potential interaction terms (none included in final models). Effects of continuous variables are presented per approximate difference between quartiles to allow their relative effects to be judged.

We used population risk attribution^23^ to quantify the contribution of selected variables to overall AMR risk, expressed as the percentage of AMR risk removed when each variable was fixed at its reference value (**Supplementary Methods**). All analyses were conducted in R.

## Results

102,955 blood specimens were taken for culture between 1 April 2015 and 31 March 2025 from 29,156 patients after their first cancer diagnostic code, of which 12,210 (11.9%) were culture positive (**Figure 1**). Excluding positive blood cultures caused by contaminants (defined in **Supplementary Methods**) left 8,154 bacteraemias. After 14-day de-duplication, there remained 5,975 bacteraemias from 4,365 patients. Of these, we included in analyses 3,141 (52.6%) bacteraemias due to Enterobacterales and 620 (10.4%) bacteraemias due to *Enterococcus faecalis/faecium* from 2,752 patients. Missing AST results in the pathogen-antimicrobial combinations studied ranged from 0.05%-0.9%, and AMR rate in these ranged from 9.2%-48.2% (**Table 1**). The three most common cancer types in the 2,752 patients included in our analyses were lymphoid/haematopoietic (630,22.9%), non-melanoma skin cancers (534,19.4%), and other cancers (520,18.9%). The median (IQR) age at the first cancer diagnostic code was 69.9 (60.5,78.8) years; 1,061 (38.6%) were female; 93 (3.4%) had non-white ethnicity, 2,205 (80.1%) white, with ethnicity missing for 454 (16.5%); the median (IQR) Charlson score and frailty scores immediately prior to cancer diagnosis were 0 (0,1.0) and 0 (0,3.2) respectively (**Table 2**). These scores had increased to 2.0 (1.0,3.0) and 9.5 (3.9,17.3) respectively at the time of bacteraemia. Reflecting studying bacteraemia from both patients with cancer and cancer survivors, the median (IQR) time since first cancer diagnostic code at the point of bacteraemia was taken was 22.4 (3.4,81.7) months, with 1577 (41.9%) taken within 12 months of diagnosis (**Table 2**).

**Table 2:** Key patient characteristics.

| Patient characteristic at first cancer diagnostic code |  | Characteristics of the included bacteraemia |
| --- | --- | --- |
| **Characteristic** | N = 2,752<sup>1</sup> | N = 3,761<sup>1</sup> |
| **Age** | 69.9 (60.5, 78.8) | 73.5 (61.7, 82.9) |
| **Sex** |  |  |
| F | 1,061 (38.6) | 2,327 (61.9) |
| M | 1,691 (61.4) | 1,434 (38.1) |
| **Ethnicity** |  |  |
| Non-white | 93 (3.4) | 154 (4.1) |
| White | 2,205 (80.1) | 2,958 (78.6) |
| Unknown | 454 (16.5) | 649 (17.3) |
| **Cancer types** |  |  |
| Breast | 159 (5.8) | 182 (4.8) |
| Colorectal | 280 (10.2) | 355 (9.4) |
| Lung | 71 (2.6) | 89 (2.4) |
| Lymphoid/Haematopoietic | 630 (22.9) | 1,011 (26.9) |
| Melanoma | 44 (1.6) | 53 (1.4) |
| Non-melanoma skin | 534 (19.4) | 687 (18.3) |
| Others | 520 (18.9) | 662 (17.6) |
| Prostate | 226 (8.2) | 285 (7.6) |
| Upper GI and hepatobiliary | 288 (10.5) | 437 (11.6) |
| **Charlson score<sup>2</sup>** | 0 (0, 1.0) | 2.0 (1.0, 3.0) |
| **Frailty score** | 0 (0, 3.2) | 9.5 (3.9, 17.3) |
| **Bacteraemia within 12 months of the first cancer diagnosis code** | — | 1577 (41.9) |
| **Months since the first cancer diagnosis code at bacteraemia diagnosis** | — | 22.4 (3.4,81.7) |
| <sup>1</sup>Median (Q1, Q3); n (%) |  |  |
| <sup>2</sup>The Charlson score was based on comorbidities immediately before the first cancer diagnostic code |  |  |

**Figure 2** summarises the multivariable models selected for each pathogen-antibiotic resistance combination (full models in **Table S2**, descriptive statistics for variables and univariable associations in **Tables S3-S9**).

**Figure 2:**
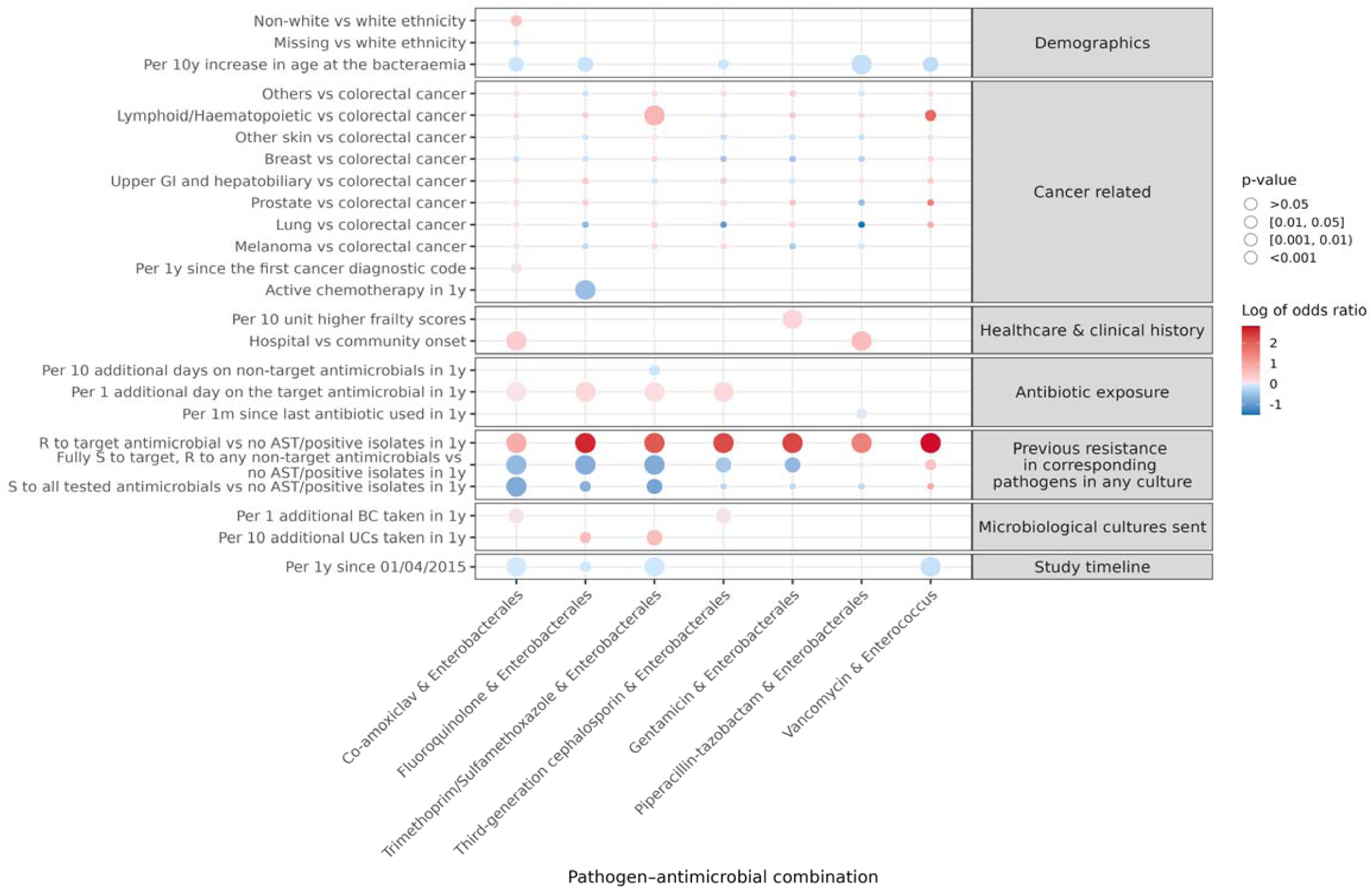
Associations between variables selected using backwards elimination and resistant vs susceptible bacteraemia for each pathogen-antimicrobial combination. Detailed results in Supplementary Tables S2-S9. Other skin cancer = Non-melanoma skin cancer, BC=blood culture, UC=urine culture.

### Associations with previous AMR

The greatest risk factor for AMR bacteraemia in patients with cancer and cancer survivors after adjustment for all other factors, was previous AMR infection in the last year (**Figure 2**). Compared with patients with no previous AST results from the corresponding pathogen in any cultures, resistance in the corresponding pathogen in any previous culture within the last year was strongly associated with increased AMR to the same antimicrobial across all pathogen-antimicrobial combinations (all p≤0.001). There were large effect sizes, with the strongest association for vancomycin-resistant *Enterococcus faecalis/faecium* (aOR= 16.94 (95% CI 6.04-47.54)), then Enterobacterales bacteraemias that were fluoroquinolone-resistant (aOR=15.23 (9.82-23.62)), gentamicin-resistant (aOR=11.17 (7.37-16.92)), third-generation cephalosporin-resistant (aOR=10.61 (7.31-15.41)), trimethoprim-sulfamethoxazole-resistant (aOR=8.84 (6.34-12.33)), piperacillin-tazobactam-resistant (aOR=4.75 (3.06-7.38)), and co-amoxiclav-resistant (aOR=2.44 (1.95-3.05)) (**Figure 3**).

**Figure 3:**
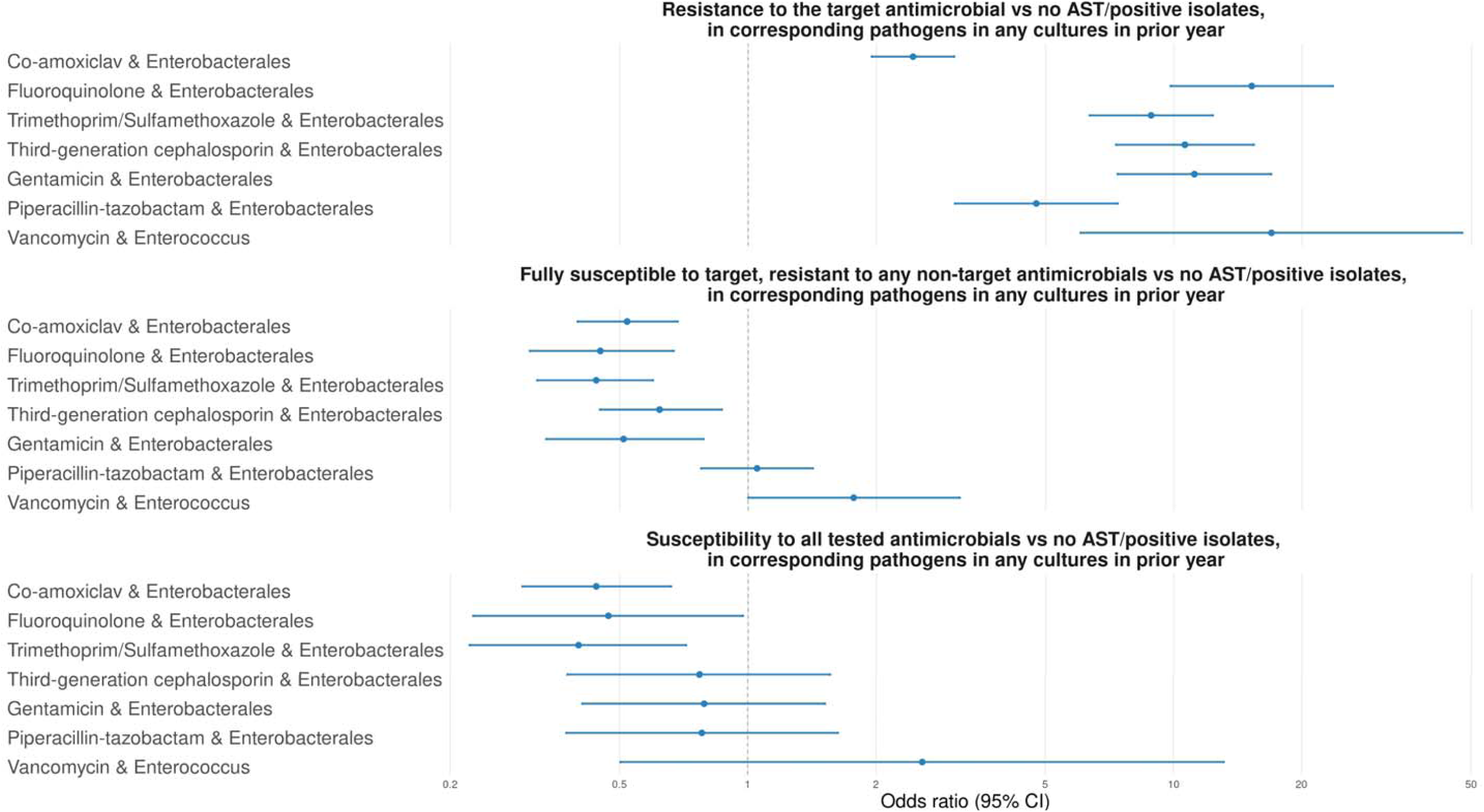
Adjusted odds ratios and 95% CIs for association between resistance/susceptibility in prior year for all pathogen-antimicrobial combinations.

Conversely, prior susceptibility to the antimicrobial of interest was associated with a reduced risk of AMR, compared to patients with no previous positive microbiology containing the pathogen of interest. In patients where all previous results for the corresponding pathogen from the last year were susceptible to the target antimicrobial, but with resistance to other antimicrobials, the odds of AMR in Enterobacterales bacteraemias were reduced relative to patients with no prior positive cultures in the last year: fluoroquinolone-resistant (aOR=0.45 (95% CI 0.31-0.67)), gentamicin-resistant (aOR= 0.51 (0.34-0.79)), third-generation cephalosporin-resistant (aOR=0.62 (0.45-0.87)), trimethoprim-sulfamethoxazole-resistant (aOR=0.44 (0.32-0.60)) and co-amoxiclav-resistant (aOR=0.52 (0.40-0.68)) (all p≤0.005).

However, previous susceptibility to vancomycin (with other resistance) was positively associated with vancomycin-resistant *Enterococcus faecalis/faecium* (aOR=1.77 (1.00, 3.14), p=0.05), with no evidence of effect on piperacillin-tazobactam-resistant Enterobacterales bacteraemia (p=0.75; hence excluded from the final model in **Figure 2**). Prior susceptibility to all tested antimicrobials in any cultures similarly reduced the odds (vs no previous AST results) of Enterobacterales bacteraemias that were resistant to trimethoprim-sulfamethoxazole (aOR=0.40 (0.22-0.72)), co-amoxiclav (aOR=0.44 (0.30-0.66)), and fluoroquinolones (aOR=0.47 (0.23-0.97)).

### Associations with previous antibiotic exposure

More days on co-amoxiclav, trimethoprim-sulfamethoxazole, fluoroquinolones, and third-generation cephalosporins in the year preceding bacteraemia were associated with higher odds of AMR to the same agent in Enterobacterales bacteraemias even after adjusting for prior detected resistance, although the effect sizes varied (aOR per day longer [95% CI]: 1.05 [1.02-1.08], 1.15 [1.07-1.24], 1.26 [1.11-1.44], and 1.21 [1.09-1.35], respectively; **Table S2**, **Figure 2**). Days on vancomycin, gentamicin, and piperacillin-tazobactam were not retained in the respective final models (p=0.09, 0.48 and 0.12 respectively adding each into the final selected model; but when added to the models had effect sizes in the same direction).

Conversely, more time on other antibiotics (i.e. not trimethoprim-sulfamethoxazole) was associated with a lower risk of trimethoprim-sulfamethoxazole-resistant Enterobacterales (aOR=0.89 (0.79, 0.99) per 10 days longer), and longer time since the last use of any antibiotic in the previous year was weakly associated with a lower risk of piperacillin-tazobactam-resistant Enterobacterales bacteraemia (aOR=0.97 (0.95-0.99) per month longer).

### Associations with cancer-related factors

Cancer type had a relatively modest and inconsistent association with AMR across pathogen-antimicrobial groups (**Figure 2**), except for haematological malignancies. Compared with colorectal cancer, lymphoid or haematopoietic malignancies were associated with higher odds of trimethoprim-sulfamethoxazole-resistant Enterobacterales bacteraemia (aOR=2.07 (95% CI, 1.40-3.06) and vancomycin-resistant *Enterococcus* bacteraemia (aOR=6.68 (1.21-36.91)). Years since the first cancer diagnostic code was associated with higher odds of third-generation-cephalosporin-resistant Enterobacterales bacteraemia (aOR=1.02 (1.00-1.04) per year longer). In contrast, having had any cancer treatment in the past year was associated with lower odds of fluoroquinolone resistant Enterobacterales (aOR=0.55 (0.39-0.77)).

### Associations with demographics

Demographic effects were generally modest across outcomes (**Figure 2**, top panel). Younger age at sample collection was independently associated with higher odds of Enterobacterales bacteraemia resistant to co-amoxiclav, fluoroquinolones, third-generation cephalosporins, and piperacillin-tazobactam, and vancomycin-resistant *Enterococcus* (adjusted odds ratios (aORs) per 10-year older 0.91 (95% CI, 0.86-0.98), 0.86 (0.77-0.96), 0.91 (0.83-0.99), 0.83 (0.76-0.91), and 0.80 (0.69-0.93) respectively) but not other pathogen-antimicrobial combinations. non-White ethnicity was associated with higher odds of co-amoxiclav-resistant Enterobacterales bacteraemia (aOR=1.74 (1.12-2.69)).

### Associations with other risk factors

Among other variables, higher frailty scores were associated with higher odds of gentamicin-resistant Enterobacterales bacteraemia (aOR=1.30 per 10 points higher (95% CI 1.12-1.50), p<0.001). Each additional blood culture taken within the previous year slightly increased the odds of co-amoxiclav-resistant and third-generation cephalosporin-resistant Enterobacterales bacteraemia (both had aOR=1.03 (1.01-1.05)): effects were stronger for urine cultures for fluoroquinolone-resistant (aOR=1.93 (1.14-3.25)) and trimethoprim-sulfamethoxazole-resistant (aOR=1.91 (1.26-2.89)) Enterobacterales bacteraemia. Similarly, hospital-onset infections were associated with co-amoxiclav-resistant (aOR=1.49 (1.23-1.82)), and piperacillin-tazobactam-resistant (aOR=1.93 (1.46-2.55)) Enterobacterales bacteraemia. The odds of resistance reduced over calendar time for co-amoxiclav-, fluoroquinolone- and trimethoprim-sulfamethoxazole-resistant Enterobacterales, and vancomycin-resistant *Enterococcus*, bacteraemias.

### Risk attribution

We evaluated how much of the total AMR was potentially attributable to each risk factor using population attributable fractions (PAF) (**Figure 4**). As expected, if we set each variable to be the reference value for the entire population then a proportion of the total AMR was removed.

**Figure 4:**
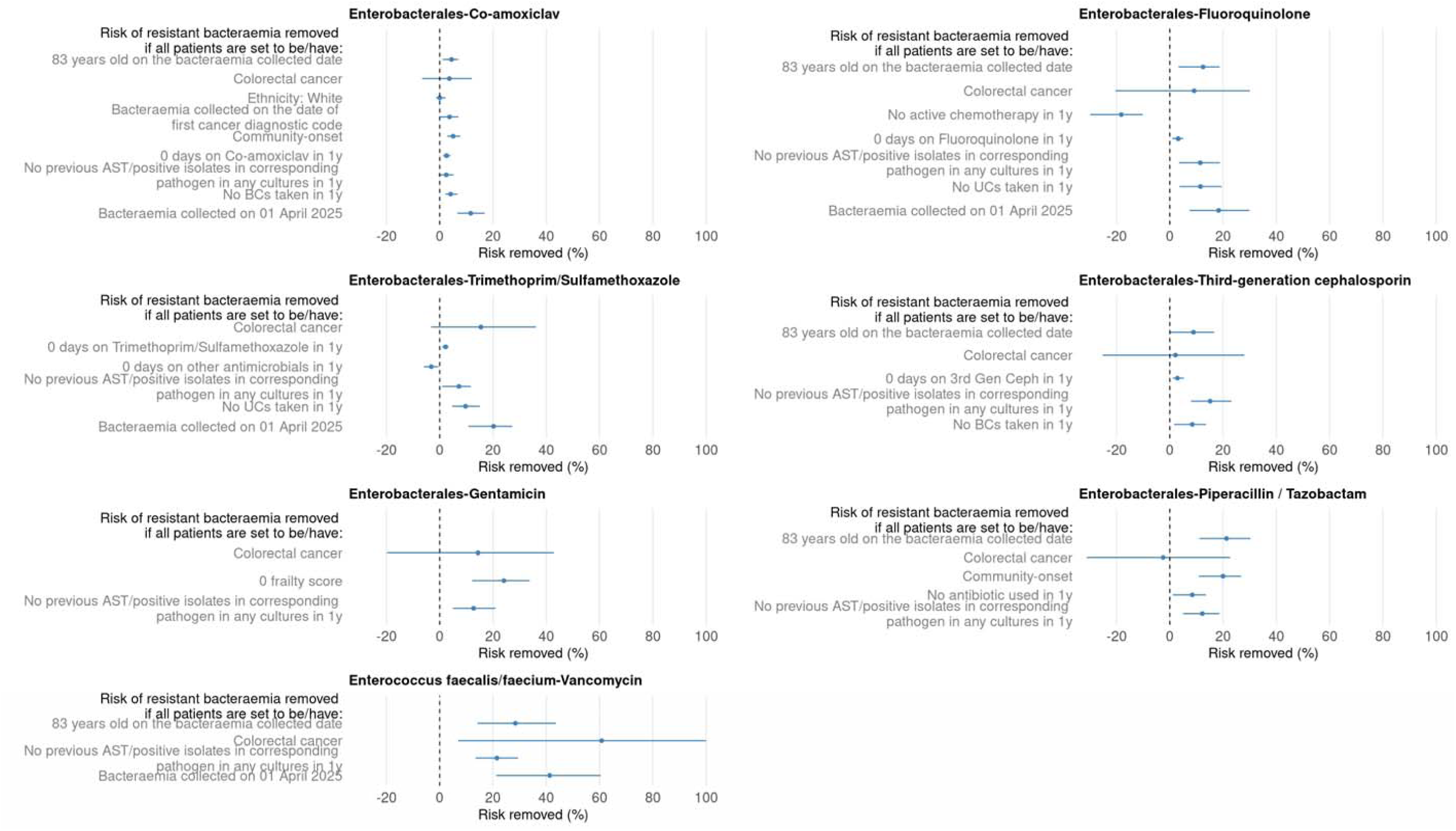
Population attributable fraction (95% CIs) assuming each variable shown could be replaced with their reference values for all patients. 83 is the third quartile of age on the blood infection date. BC=blood culture, UC=urine culture.

Assuming all patients had no prior positive cultures for corresponding pathogen reduced AMR by 2.5%-21.5% (**Figure 4**), with the largest contributions from this variable for vancomycin-resistant *Enterococcus* bacteraemia (21.5%), then third-generation cephalosporin- (15.2%), gentamicin- (12.7%), piperacillin-tazobactam- (12.2%), fluoroquinolone (11.4%), trimethoprim-sulfamethoxazole- (7.2%), and co-amoxiclav- (2.5%) resistant Enterobacterales. In contrast, the PAFs assuming zero days on the target antimicrobials in the last year were much smaller, ranging from only 2.2%-3.1% (albeit while holding rates of previously detected AMR constant).

If the index time for the entire population was set to April 2025, the estimated overall risk removed ranged from 11.6%-41.3% across pathogen-antimicrobial combinations reflecting local reductions in AMR over time. Assuming all patients were aged 83 years (the third quartile) reduced AMR by 4.4%-28.4% for the whole population (due to higher risks at younger ages). Although non-white ethnicity was associated with higher odds of resistance (aOR=1.74), because only a minority of patients were of non-white ethnicity, assigning all patients to white ethnicity resulted in a minimal PAF of 0.02%. This reflects the fact that PAF depends on both the strength of association and the prevalence of exposure: factors with strong effects but low prevalence may contribute little to the overall population risk.

Assuming all patients had received cancer treatment in the last year resulted in an 18.2% reduction in overall risk for fluoroquinolone-resistant Enterobacterales. Finally, assigning all patients to have colorectal cancer, to assess the impact of cancer type, was associated with a 60.8% reduction vancomycin-resistant *Enterococcus* bacteraemia reflecting particularly the association with haematological malignancy; estimated impact of cancer type for other pathogen-antimicrobial combinations were uncertain with large confidence intervals.

## Discussion

We show that in patients with cancer and cancer survivors who develop a bacteraemia, prior AMR and prior antibiotic exposure were important determinants of AMR risk. Increased risks were also seen in younger patients and those with haematological malignancies. Lower risks of AMR were seen for Enterobacterales with a recent antimicrobial susceptible isolate.

Our findings align broadly with other studies from different population groups, e.g., those showing resistance in the last year in Gram-negative isolates to a specific antibiotic is associated with future resistance to that same drug in bloodstream infections^11^ while previous susceptive urine cultures within 2 years were effective for identifying future susceptibility to agents such as trimethoprim-sulfamethoxazole, gentamicin, piperacillin-tazobactam, and fluoroquinolones in urine cultures ^24^. In this current study, across a broader range of ‘bug-drug’ combinations and specimen types, we found that previous resistance within the past year was a major driver of resistant bacteraemias, whereas prior susceptibility was positively associated with antimicrobial susceptibility in Enterobacterales bacteraemias but interestingly not *Enterococcus* bacteraemias, potentially reflecting less stability in Enterococcal carriage strains vs. Enterobacterales. Again consistent with previous work^25, 26^, we showed prior exposure to the target antimicrobial increased AMR risks, particularly for third-generation cephalosporins, co-amoxiclav, fluoroquinolones, and trimethoprim/sulfamethoxazole, although effects were modest compared with the effect of prior resistance/susceptibility. Interestingly, exposure to antibiotics other than trimethoprim-sulfamethoxazole was positively associated with lower risks of trimethoprim-sulfamethoxazole-resistant Enterobacterales, possibly due to elimination of resistant strains, although the reasons for this remain unclear.

Patients undergoing hematopoietic stem cell transplantation have previously been shown to be at particularly high risk for vancomycin-resistant *Enterococcus* infections^27, 28^, which is consistent with our finding that compared with colorectal cancers, haematological malignancies were associated with higher odds of vancomycin resistance. We also observed an association between lymphoid/haematopoietic malignancies and Enterobacterales resistant to trimethoprim/sulfamethoxazole, possibly due to the widespread use of this agent for prophylaxis in haematology patients^29^.

Previous studies have suggested that cancer chemotherapy increases AMR risk through prolonged antibiotic use during treatment^30^ and via gut microbiota disruption caused by chemotherapy^16^. In contrast, our study found that recent cancer treatment was negatively associated with fluoroquinolone resistance in Enterobacterales bacteraemias, with no evidence of effect for other pathogen-drug combinations. In part this may be because we adjust for other factors more completely than earlier studies, including antibiotic exposure. It may also represent healthier patients being more likely receive chemotherapy and to continue it.

Patients with hospital-onset infections also showed increased risk of resistance to co-amoxiclav, and piperacillin-tazobactam in Enterobacterales, potentially reflecting acquisition of these resistance patterns specifically from the hospital environment and consistent with previous work showing hospital-onset infection as a risk factor for AMR^31^.

### Limitations

This study has some limitations. First, linkage to cancer registry data was not available, and therefore cancers were identified using diagnostic codes in hospital data. Compared to linked data sources, the sensitivity of hospital data to capture cancer is generally in the range of 70-90%, with the exception of 55% for malignant melanoma. Specificity, however, it has been shown to be very high, with positive predictive values above 90% for the most common cancers^32^. As diagnoses were inferred from the first recorded code, the true diagnosis or its timing may be inaccurate and lead to dilution bias in estimated effects of time since diagnosis, although previous studies suggest time differences between coded diagnoses and registry data are typically <30 days^32^. Additionally, our study was conducted using data only from Oxfordshire (1% of the UK population) and we only considered resistance in blood cultures as outcome, which may limit generalisability. Finally, our data did not include antibiotics prescribed in primary care limiting ascertainment of prior antibiotic exposure.

Overall, our study highlights the importance of AMR in patients with cancer and cancer survivors. Most cancer research is focused on cancer itself, with 35-fold more publications than on priority bacteria^33^. We show how AMR undermines cancer treatment and outcomes for cancer survivors, emphasising the need to also focus investment in antibacterial innovation. Our findings emphasise the need to consider increased risks of AMR when giving empirical treatment, and the need to develop better interventions to reduce AMR in this important patient group.

## Author contributions

D.W.E. and A.S.W. designed the study. F.H. and J.W. carried out the data analysis. All authors contributed to drafting the manuscript and to its critical review and revision.

## Funding

This project was funded by the National Institute for Health Protection (NIHR) Health Protection Research Unit (F.H., A.S.W., D.E., J.W., R.C.) in Healthcare Associated Infections and Antimicrobial Resistance at Oxford University in partnership with the UK Health Security Agency (UKHSA) (NIHR207397) and supported by the NIHR Biomedical Research Centre (A.S.W), Oxford. The views expressed are those of the authors and not necessarily those of the NHS, the NIHR, the Department of Health and Social Care or UKHSA. ASW is an NIHR Senior Investigator.

## Data Availability

Due to the sensitive nature of the patient-level electronic health records and the terms of the ethical approval, the raw dataset cannot be made publicly available or shared directly by the authors. De-identified data may be made available to researchers upon reasonable request, subject to a formal application to the IORD Research Committee and the execution of a data sharing agreement. Further information on the application process and data access policies can be found at the IORD website or by contacting the database administrators at the Big Data Institute, University of Oxford.

## Acknowledgements

This work uses data provided by patients and collected by the UK’s National Health Service as part of their care and support. We thank all the people of Oxfordshire who contribute to the Infections in Oxfordshire Research Database.

Research Database Team: L Butcher, H Boseley, C Crichton, DW Crook, J Davies, D Eyre, R Harrington, G Hayward, K Jeffery, F Kemp, E Morris, TEA Peto, D Prieto-Alhambra, TP Quan, R Shackell, B Shine, AS Walker, K Woods.

Patient and Public Panel: M Ahmed, G Blower, J Hopkins, R Mandunya, S Markham.

For the purpose of Open Access, the author has applied a CC BY public copyright licence to any Author Accepted Manuscript (AAM) version arising.

## Declaration of Interest Statement

All authors declare no conflicts of interest.

## Notes

### Competing Interest Statement

The authors have declared no competing interest.

### Author Declarations

We used data from the Infections in Oxfordshire Research Database (IORD). IORD has approvals from the National Research Ethics Service South Central Oxford C Research Ethics Committee (24/SC/0241), the Health Research Authority, and the Confidentiality Advisory Group (19/CAG/0144) as a research database not requiring individual patient consent.

## References

1. Organisation WH. Global action plan on antimicrobial resistance. Global action plan on antimicrobial resistance (who int). 2015.

2. Murray CJ, Ikuta KS, Sharara F, et al. Global burden of bacterial antimicrobial resistance in 2019: a systematic analysis. The lancet. 2022;399(10325):629–655.

3. Mahase E. Antibiotic resistant infection deaths increasing in England, despite lower NHS antibiotic use. British Medical Journal Publishing Group 2025.

4. Zembower TR. Epidemiology of infections in cancer patients. Infectious complications in cancer patients. 2014:43–89.

5. Rolston KV. Infections in cancer patients with solid tumors: a review. Infectious diseases and therapy. 2017;6(1):69–83.

6. Gupta V, Satlin MJ, Kalvin CY, et al. Incidence and prevalence of antimicrobial resistance in outpatients with cancer: a multicentre, retrospective, cohort study. The Lancet Oncology. 2025;26(5):620–628.

7. Teillant A, Gandra S, Barter D, et al. Potential burden of antibiotic resistance on surgery and cancer chemotherapy antibiotic prophylaxis in the USA: a literature review and modelling study. The Lancet infectious diseases. 2015;15(12):1429–1437.

8. English surveillance programme for antimicrobial utilisation and resistance (ESPAUR) Report 2024 to 2025. UK Health Security Agency (UKHSA) 2025.

9. Larramendy S, Deglaire V, Dusollier P, et al. Risk factors of extended-spectrum beta-lactamases-producing Escherichia coli community acquired urinary tract infections: a systematic review. Infection and Drug Resistance. 2020:3945–3955.

10. Halldórsdóttir AM, Hrafnkelsson B, Einarsdóttir K, et al. Prevalence and risk factors of extended-spectrum beta-lactamase producing E. coli causing urinary tract infections in Iceland during 2012–2021. European Journal of Clinical Microbiology & Infectious Diseases. 2024;43(9):1689–1697.

11. MacFadden D, Coburn B, Shah N, et al. Utility of prior cultures in predicting antibiotic resistance of bloodstream infections due to Gram-negative pathogens: a multicentre observational cohort study. Clinical Microbiology and Infection. 2018;24(5):493–499.

12. Kahlmeter G, Brown D, Goldstein F, et al. European Committee on Antimicrobial Susceptibility Testing (EUCAST) technical notes on antimicrobial susceptibility testing. Wiley Online Library 2006:501–503.

13. Yoon CH, Bartlett S, Stoesser N, et al. Mortality risks associated with empirical antibiotic activity in Escherichia coli bacteraemia: an analysis of electronic health records. Journal of Antimicrobial Chemotherapy. 2022;77(9):2536–2545.

14. Yuan K, Luk A, Wei J, et al. Machine learning and clinician predictions of antibiotic resistance in Enterobacterales bloodstream infections. Journal of Infection. 2025;90(2):106388.

15. Haeusler GM, Mechinaud F, Daley AJ, et al. Antibiotic-resistant Gram-negative bacteremia in pediatric oncology patients—risk factors and outcomes. The Pediatric infectious disease journal. 2013;32(7):723–726.

16. Nanayakkara AK, Boucher HW, Fowler Jr VG, et al. Antibiotic resistance in the patient with cancer: Escalating challenges and paths forward. CA: a cancer journal for clinicians. 2021;71(6):488–504.

17. Mahadevan P, Harley M, Fordyce S, et al. Completeness and representativeness of small area socioeconomic data linked with the UK Clinical Practice Research Datalink (CPRD). J Epidemiol Community Health. 2022;76(10):880–886.

18. Charlson ME, Pompei P, Ales KL, et al. A new method of classifying prognostic comorbidity in longitudinal studies: development and validation. Journal of chronic diseases. 1987;40(5):373–383.

19. Quan H, Sundararajan V, Halfon P, et al. Coding algorithms for defining comorbidities in ICD-9-CM and ICD-10 administrative data. Medical care. 2005;43(11):1130–1139.

20. Gilbert T, Neuburger J, Kraindler J, et al. Development and validation of a Hospital Frailty Risk Score focusing on older people in acute care settings using electronic hospital records: an observational study. The Lancet. 2018;391(10132):1775–1782.

21. Zeileis A. Object-oriented computation of sandwich estimators. Journal of statistical software. 2006;16:1–16.

22. Lee DK. Alternatives to P value: confidence interval and effect size. Korean journal of anesthesiology. 2016;69(6):555.

23. Greenland S, Drescher K. Maximum likelihood estimation of the attributable fraction from logistic models. Biometrics. 1993:865–872.

24. Valentine-King MA, Trautner BW, Zoorob RJ, et al. Predicting antibiotic susceptibility among patients with recurrent urinary tract infection using a prior culture. The Journal of Urology. 2024;211(1):144–152.

25. Costelloe C, Metcalfe C, Lovering A, et al. Effect of antibiotic prescribing in primary care on antimicrobial resistance in individual patients: systematic review and meta-analysis. Bmj. 2010;340.

26. Baraz A, Chowers M, Nevo D, et al. The time-varying association between previous antibiotic use and antibiotic resistance. Clinical Microbiology and Infection. 2023;29(3):390. e391–390. e394.

27. Ford CD, Gazdik MA, Lopansri BK, et al. Vancomycin-resistant Enterococcus colonization and bacteremia and hematopoietic stem cell transplantation outcomes. Biology of Blood and Marrow Transplantation. 2017;23(2):340–346.

28. So M. Vancomycin-resistant Enterococcus in hematology-oncology patients: a review on colonization, screening, infections, resistance, and antimicrobial stewardship. Current Treatment Options in Infectious Diseases. 2020;12(3):285–295.

29. Ward T, Thomas R, Fye C, et al. Trimethoprim-sulfamethoxazole prophylaxis in granulocytopenic patients with acute leukemia: evaluation of serum antibiotic levels in a randomized, double-blind, placebo-controlled Department of Veterans Affairs Cooperative Study. Clinical infectious diseases. 1993;17(3):323–332.

30. Kenneth MJ, Wu C-C, Fang C-Y, et al. Exploring the Impact of Chemotherapy on the Emergence of Antibiotic Resistance in the Gut Microbiota of Colorectal Cancer Patients. Antibiotics. 2025;14(3):264.

31. Abban MK AE, Mosi L, Isawumi A. The burden of hospital acquired infections and antimicrobial resistance. Heliyon. 2023;9(10).

32. Strongman H, Williams R, Bhaskaran K. What are the implications of using individual and combined sources of routinely collected data to identify and characterise incident site-specific cancers? a concordance and validation study using linked English electronic health records data. BMJ open. 2020;10(8):e037719.

33. Alliance AI. Leaving the lab: tracking the decline in AMR R&D professionals. AMR Industry Alliance. 2024.

