## Supplementary Material for "Risk Factors for Antimicrobial Resistance in Cancer Patients and Cancer Survivors: An Electronic Health Record Study"

**RISK FACTORS FOR ANTIMICROBIAL RESISTANCE IN PATIENTS WITH CANCER OR A HISTORY OF CANCER: AN ELECTRONIC HEALTH RECORD STUDY: Supplementary appendix**

### Supplementary methods

#### Variables and definitions

Cancer types were defined according to the following ICD-10 codes: C00–C26 (malignant neoplasms of the lip, oral cavity, pharynx, gastrointestinal tract, and intra-abdominal organs), C30–C39 (malignant neoplasms of respiratory and intrathoracic organs), C40–C41 (malignant neoplasms of bone and articular cartilage), C43–C44 (melanoma and other malignant neoplasms of skin), C45–C49 (malignant neoplasms of mesothelial and soft tissue), C50 (malignant neoplasm of breast), C51–C58 (malignant neoplasms of female genital organs), C60–C63 (malignant neoplasms of male genital organs), C64–C68 (malignant neoplasms of urinary tract), C69–C72 (malignant neoplasms of the eye, brain, and other parts of the central nervous system), C73–C75 (malignant neoplasms of the thyroid and other endocrine glands), C76–C80 (malignant neoplasms of ill-defined, secondary, and unspecified sites), C81–C96 (malignant neoplasms, stated or presumed to be primary, of lymphoid, haematopoietic, and related tissue), and C97 (malignant neoplasms of independent [primary] multiple sites). The cancer types were then grouped to breast cancer (C50), prostate cancer (C61), colorectal cancer (C18-20), upper gastrointestinal and hepatobiliary cancer (C15-17, C22-25), melanoma cancer (C43), non-melanoma skin cancers (C44), lung cancer (C34), lymphoid/haematopoietic cancer (C81-88, C90-96), and other cancers for the rest.

Contaminants were defined by the name of the isolated organism containing ‘COAGULASE NEGATIVE STAPH’, ‘CoNS’, ‘STAPH’ (without other speciation), ‘PROPIONIBACTERIUM’, ’DIPHTHEROIDS’, ’MICROCOCCUS’, ‘BACILLUS’, ‘AEROBIC SPORE BEARER’, ‘CORYNEBACTERIUM’. For Charlson and frailty scores, ICD-10 codes were derived from prior consultant episodes finishing strictly before the index consultant episode containing the sample collection date using a 1 year lookback.

We included variables derived from microbiology samples from both blood and non-blood specimens. Non-blood cultures contained: urine culture, surface swab culture, pus microscopy/culture, Methicillin-resistant Staphylococcus aureus (MRSA) screen, respiratory culture and microscopy, sterile site culture, genital culture, carbapenemase-producing Enterobacterales (CPE) screen, and line tip culture. To account for previous antimicrobial resistance (AMR) exposure, we considered whether within one year before the sample collection date each pathogen-antimicrobial group showed: any resistance in any (including both blood and non-blood) cultures to the corresponding antimicrobial, any resistance in any cultures to other antimicrobials than the corresponding one (equivalent to fully susceptible to the target and resistant to any non-target antimicrobials in any cultures), fully susceptible to all tested antimicrobials in prior AST, and no prior AST or positive isolates from the corresponding pathogens.

We defined the number of days admitted to OUH hospitals, days in the intensive care unit (ICU), and days on (specific or any) antibiotics during the past year as the total number of unique days of hospital admission, ICU stay, and antibiotic exposure within one year prior to the sample collection date, regardless of whether these days were consecutive. To reflect the long half-lives and effects of chemotherapy agents, days on active chemotherapy in the past year was defined as the total number of days within one year before the sample collection date plus an additional 14 days for each discrete cycle of active chemotherapy (a new cycle was defined by a chemotherapy course starting >90 days from the last course).

All continuous variables were truncated at the 5th and 95th percentiles to minimise the influence of outliers on the model.

##### Non-linearity

Non-linearity was assessed in univariable logistic regression models using restricted cubic splines. Splines with one to four internal knots were considered for all continuous variables. After truncating variable values at the 5th and 95th percentiles, internal knots were placed at evenly spaced percentiles of the range of values of each variable, and boundary knots were set as the 10th and 90th percentiles of the range. The optimal degree of non-linearity was selected based on the Bayesian Information Criterion (BIC), with a difference greater than 2 interpreted as positive evidence favouring the model with the lower BIC. In multivariable models with all potential risk factors, spline terms were re-evaluated using BIC to prevent overfitting, and linear terms replaced spline terms when the model if the linear terms had a lower BIC.

###### Population risk attribution

To estimate the percentage of risk attributable to each variable $X_{j}$ when fixed at its reference value $r_{j}$ across the entire population, we used the following formula^1^:

$$\text{PAF}=\frac{\sum_{i} \left( p_{i}\left( \text{AMR} \right)-\tilde{p_{i}}\left( \text{AMR} | X_{j}=r_{j} \right) \right)}{\sum_{i} p_{i}\left( \text{AMR} \right)} ,$$

where $p_{i}$(AMR) represents the predicted probability of AMR for individual $i$ under observed exposure values based on the final multivariable model and $\tilde{p_{i}}\left( \text{AMR} | X_{j}=r_{j} \right)$ denotes the predicted probability for the same individual if the variable $X_{j}$ were set to its reference value $r_{j}$. PAF would be negative if $\sum_{i} p_{i}\left( \text{AMR} \right) < \sum_{i} \tilde{p_{i}}\left( \text{AMR} | X_{j}=r_{j} \right)$, indicating that the overall risk would increase if the variable $X_{j}$ was set to be the reference value $r_{j}$ for entire population, i.e. the factor is associated with a protective effect.

The 95% confidence intervals for the PAF were estimated using the percentile bootstrap method^2^. Specifically, 100 bootstrap samples were drawn with replacement from the original dataset, and the PAF was recalculated for each sample. The 2.5th and 97.5th percentiles of the resulting bootstrap distribution were then taken as the lower and upper bounds of the 95% CI, respectively.

###### Supplementary Tables

| Category | Variables |
| --- | --- |
| Demographics and comorbidity | 1. Age (continuous) 2. Sex: Female, Male (categorical) 3. Ethnicity: White, Non-white, Missing (categorical) 4. Deprivation percentile (continuous)   5. Charlson score (continuous) |
| Cancer related | 6. Cancer type at the first diagnostic code: breast, prostate, colorectal, upper gastrointestinal and hepatobiliary, melanoma, non-melanoma skin, lung, lymphoid/haematopoietic, other cancers (categorical) 7. Time since the first cancer diagnostic code (continuous)  8. Cancer treatment (including chemotherapy and radiotherapy) in the past year (binary)  9. Total days on cancer treatment (continuous)  10. Number of neutrophil tests taken in the past year (continuous) |
| Healthcare and clinical history | 11. Frailty score (continuous) 12. Any positive catheter specimens sent for urine cultures in the last year (binary) 13. Days in hospitals in the last year (continuous) 14. Days in ICU in the last year (continuous)  15. Blood infection onset: Hospital (>48h after admission and before discharge), Community (categorical) |
| Antibiotics exposure | For predefined pathogen-antimicrobial combinations in **Table 1**:  16. Number of days on the target antimicrobial prescribed by hospitals in the last year (continuous)  17. Number of days on the non-target antimicrobial prescribed by hospitals in the last year (continuous)  18. Time since the last antibiotic used in the last year (continuous) |
| Previous AMR | 19. Previous resistance status in the past year for predefined pathogen-antimicrobial combinations in **Table 1**: in the corresponding pathogen in any cultures,  No previous AST result/positive isolates, Having had any isolate with resistance to the target antimicrobial, Having had any isolate with resistance but only to non-target antimicrobials (equivalent to having been fully susceptible to the target and resistant to any non-target antimicrobials), Fully susceptible to all tested antimicrobials (categorical) |
| Microbiological cultures sent | 20. Number of urine cultures taken in the past year (continuous)  21. Number of blood cultures take in the past year (continuous) |
| Study Timeline | 22. Calendar time from 1 April 2015 (continuous) |

###### Table S1: List of variables considered.

|  | Multivariable (adjusted) with Backward Selection Results | | | | | | | | | | | | | | |
| --- | --- | --- | --- | --- | --- | --- | --- | --- | --- | --- | --- | --- | --- | --- | --- |
|  | | **Co-amoxiclav in Enterobacterales** | | **Fluoroquinolone in Enterobacterales** | | **Co-Trimethoprim in Enterobacterales** | | **3rd generation-cephalosporin in Enterobacterales** | | **Gentamicin in Enterobacterales** | | **Piperacillin-tazobactam in Enterobacterales** | | **Vancomycin in Enterococcus faecalis/faecium** | |
| **Variable** | | **OR (95% CI)** | **p-value** | **OR (95% CI)** | **p-value** | **OR (95% CI)** | **p-value** | **OR (95% CI)** | **p-value** | **OR (95% CI)** | **p-value** | **OR (95% CI)** | **p-value** | **OR (95% CI)** | **p-value** |
| **Demographics Characteristics:** | |  |  |  |  |  |  |  |  |  |  |  |  |  |  |
| **Age at Bloodstream Infection, per 10 years** | | 0.91 (0.86,0.98) | **0.008** | 0.86 (0.77,0.96) | **0.005** |  |  | 0.91 (0.83,0.99) | **0.05** |  |  | 0.83 (0.76,0.91) | **<0.001** | 0.80 (0.69,0.93) | **0.002** |
| **Ethnicity: Non-white vs White** | | 1.74 (1.12,2.69) | **0.01** |  |  |  |  |  |  |  |  |  |  |  |  |
| **Ethnicity: Missing vs White** | | 0.88 (0.70,1.12) | 0.30 |  |  |  |  |  |  |  |  |  |  |  |  |
| **Cancer Types and Cancer related:** | |  |  |  |  |  |  |  |  |  |  |  |  |  |  |
| Others vs Colorectal Cancer | | 1.10 (0.82,1.48) | 0.52 | 0.91 (0.54,1.54) | 0.73 | 1.15 (0.75,1.75) | 0.52 | 1.20 (0.75,1.94) | 0.45 | 1.45 (0.81,2.59) | 0.21 | 0.95 (0.58,1.56) | 0.85 | 1.13 (0.18,7.25) | 0.89 |
| Lymphoid/Haematopoietic vs Colorectal Cancer | | 1.18 (0.87,1.59) | 0.28 | 1.48 (0.91,2.41) | 0.11 | 2.07 (1.40,3.06) | **<0.001** | 0.97 (0.60,1.57) | 0.89 | 1.56 (0.87,2.77) | 0.13 | 1.18 (0.75,1.86) | 0.48 | 6.68 (1.21,36.91) | **0.03** |
| Non-melanoma skin vs Colorectal Cancer | | 1.05 (0.78,1.41) | 0.74 | 0.91 (0.53,1.55) | 0.72 | 1.06 (0.70,1.61) | 0.78 | 0.85 (0.51,1.39) | 0.51 | 0.90 (0.47,1.69) | 0.74 | 0.87 (0.52,1.45) | 0.59 | 1.02 (0.13,8.09) | 0.99 |
| Breast vs Colorectal Cancer | | 0.90 (0.60,1.33) | 0.59 | 0.86 (0.43,1.70) | 0.66 | 1.29 (0.74,2.23) | 0.37 | 0.61 (0.30,1.25) | 0.18 | 0.60 (0.22,1.66) | 0.33 | 0.73 (0.34,1.54) | 0.41 | 1.26 (0.15,10.36) | 0.83 |
| Upper GI and hepatobiliary vs Colorectal Cancer | | 1.12 (0.81,1.54) | 0.49 | 1.54 (0.91,2.61) | 0.11 | 0.94 (0.59,1.50) | 0.79 | 1.48 (0.88,2.49) | 0.14 | 0.95 (0.46,1.97) | 0.89 | 1.06 (0.61,1.83) | 0.84 | 1.54 (0.25,9.71) | 0.64 |
| Prostate vs Colorectal Cancer | | 1.15 (0.81,1.64) | 0.44 | 1.41 (0.79,2.54) | 0.25 | 1.03 (0.63,1.66) | 0.91 | 1.23 (0.69,2.21) | 0.48 | 1.71 (0.90,3.23) | 0.09 | 0.55 (0.27,1.15) | 0.11 | 5.26 (0.75,37.03) | 0.09 |
| Lung vs Colorectal Cancer | | 1.04 (0.61,1.75) | 0.89 | 0.52 (0.18,1.47) | 0.22 | 1.31 (0.66,2.60) | 0.44 | 0.34 (0.09,1.31) | 0.12 | 1.28 (0.46,3.58) | 0.64 | 0.22 (0.03,1.74) | 0.15 | 2.36 (0.22,25.58) | 0.48 |
| Melanoma vs Colorectal Cancer | | 1.03 (0.54,1.94) | 0.92 | 0.83 (0.24,2.93) | 0.77 | 1.15 (0.50,2.64) | 0.74 | 1.28 (0.45,3.11) | 0.62 | 0.65 (0.14,2.92) | 0.57 | 0.92 (0.29,2.96) | 0.89 |  |  |
| **Active Chemotherapy in Last Year** | |  |  | 0.55 (0.39,0.77) | **<0.001** |  |  |  |  |  |  |  |  |  |  |
| **Years since the First Cancer Diagnostic Code, per year** | | 1.02 (1.00,1.04) | **<0.001** |  |  |  |  |  |  |  |  |  |  |  |  |
| **Healthcare Utilisation & Clinical History:** | |  |  |  |  |  |  |  |  |  |  |  |  |  |  |
| **Blood Infection Onset** | |  |  |  |  |  |  |  |  |  |  |  |  |  |  |
| Hospital vs Community | | 1.49 (1.23,1.82) | **<0.001** |  |  |  |  |  |  |  |  | 1.93 (1.46,2.55) | **<0.001** |  |  |
| **1-year-look-back Frailty Score, per 10 scores** | |  |  |  |  |  |  |  |  | 1.30 (1.12,1.50) | **<0.001** |  |  |  |  |
| **Antibiotics Exposure:** | |  |  |  |  |  |  |  |  |  |  |  |  |  |  |
| **Days on Non-target Antibiotics in Last Year, per 10 days** | |  |  |  |  | 0.89 (0.79,0.99) | **0.04** |  |  |  |  |  |  |  |  |
| **Days on Tested Antibiotics in Last Year, per 1 day** | | 1.05 (1.02,1.08) | **<0.001** | 1.26 (1.11,1.44) | **<0.001** | 1.15 (1.07,1.24) | **<0.001** | 1.21 (1.09,1.35) | **<0.001** |  |  |  |  |  |  |
| **Days Since Last Antibiotic Used in Last Year, per 30 days** | |  |  |  |  |  |  |  |  |  |  | 0.97 (0.95,0.99) | **0.01** |  |  |
| **Previous Resistance:** | |  |  |  |  |  |  |  |  |  |  |  |  |  |  |
| **Previous Resistance in Any Culture in the Last Year** | |  |  |  |  |  |  |  |  |  |  |  |  |  |  |
| No Previous AST Result/Positive Isolates, from Corresponding Pathogens | | — |  | — |  | — |  | — |  | — |  | — |  | — |  |
| Resistance to Target Antimicrobials in Corresponding Pathogens | | 2.44 (1.95,3.05) | **<0.001** | 15.23 (9.82,23.62) | **<0.001** | 8.84 (6.34,12.33) | **<0.001** | 10.61 (7.31,15.41) | **<0.001** | 11.17 (7.37,16.92) | **<0.001** | 4.75 (3.06,7.38) | **<0.001** | 16.94 (6.04,47.54) | **<0.001** |
| Resistance Only to Non-target Antimicrobials in Corresponding Pathogens¹ | | 0.52 (0.40,0.68) | **<0.001** | 0.45 (0.31,0.67) | **<0.001** | 0.44 (0.32,0.60) | **<0.001** | 0.62 (0.45,0.87) | **0.005** | 0.51 (0.34,0.79) | **0.002** | 1.05 (0.78,1.42) | 0.75 | 1.77 (1.00,3.14) | **0.05** |
| Fully Susceptible to All Tested Antimicrobials in Corresponding Pathogens | | 0.44 (0.30,0.66) | **<0.001** | 0.47 (0.23,0.97) | **0.005** | 0.40 (0.22,0.72) | **0.002** | 0.77 (0.38,1.56) | 0.46 | 0.79 (0.41,1.52) | 0.48 | 0.78 (0.37,1.63) | 0.51 | 2.57 (0.50,13.13) | 0.26 |
| **Testings and Infection Source:** | |  |  |  |  |  |  |  |  |  |  |  |  |  |  |
| **Number of Blood Samples Taken in Last Year, per 1 sample** | | 1.03 (1.01,1.05) | **0.003** |  |  |  |  | 1.03 (1.01,1.05) | **0.003** |  |  |  |  |  |  |
| **Number of Urine Samples Taken in Last Year, per 10 cultures** | |  |  | 1.93 (1.14,3.25) | **0.01** | 1.91 (1.26,2.89) | **0.002** |  |  |  |  |  |  |  |  |
| **Study Timeline:** | |  |  |  |  |  |  |  |  |  |  |  |  |  |  |
| **Calendar Days of Collection Date Since 01/04/2015, per year** | | 0.95 (0.92,0.97) | **<0.001** | 0.94 (0.90,0.98) | **0.01** | 0.93 (0.90,0.97) | **<0.001** |  |  |  |  |  |  | 0.85 (0.77,0.93) | **<0.001** |
| Abbreviations: CI = Confidence Interval, OR = Odds Ratio | | | | | | | | | | | | | | | |
| ¹Equivalent to fully susceptible to the target antimicrobial and resistant to any non-target antimicrobials. | | | | | | | | | | | | | | | |

Table S2: Final selected logistic regression models for associations with AMR in different pathogen and antimicrobial combinations.

| Co-amoxiclav in Enterobacterales positive blood cultures (N=3,120) | | | | | | |
| --- | --- | --- | --- | --- | --- | --- |
|  | **Susceptible N = 1,617**¹ | **Resistant N = 1,503**¹ | **Univariable (unadjusted)** | | **Multivariable (adjusted) with Backward Selection** | |
| **Variable** |  |  | **OR (95% CI)**² | **p-value** | **OR (95% CI)**² | **p-value** |
| **Demographics Characteristics and Cancer Related:** |  |  |  |  |  |  |
| **Age Group at Bloodstream Infection** |  |  |  |  |  |  |
| 60-80 | 723 (23.2) | 690 (22.1) | — |  |  |  |
| <40 | 35 (1.1) | 83 (2.7) | 2.46 (1.6, 3.78) | **<0.001** |  |  |
| 40-60 | 206 (6.6) | 294 (9.4) | 1.47 (1.16, 1.85) | **0.001** |  |  |
| 80+ | 653 (20.9) | 436 (14.0) | 0.72 (0.61,0.85) | **<0.001** |  |  |
| **Age at Bloodstream Infection, per 10 years** | 76.9 (66.9,85.0) | 72.0 (60.0,81.6) | 0.81 (0.77, 0.85) | **<0.001** | 0.91 (0.86, 0.98) | **0.008** |
| **Sex** |  |  |  |  |  |  |
| M | 993 (31.8) | 919 (29.5) | — |  | — |  |
| F | 624 (20.0) | 584 (18.7) | 1 (0.85,1.18) | 0.96 |  |  |
| **Ethnicity** |  |  |  |  |  |  |
| White | 1,322 (42.4) | 1,162 (37.2) | — |  | — |  |
| Non-white | 41 (1.3) | 87 (2.8) | 2.46 (1.59,3.81) | **<0.001** | 1.74 (1.12,2.69) | **0.01** |
| Missing | 254 (8.1) | 254 (8.1) | 1.12 (0.91,1.39) | 0.28 | 0.88 (0.70,1.12) | 0.30 |
| **Cancer Group** |  |  |  |  |  |  |
| Colorectal | 170 (5.4) | 141 (4.5) | — |  | — |  |
| Others | 290 (9.3) | 272 (8.7) | 1.12 (0.82,1.53) | 0.48 | 1.10 (0.82,1.48) | 0.52 |
| Lymphoid/Haematopoietic | 298 (9.6) | 428 (13.7) | 1.74 (1.28,2.35) | **<0.001** | 1.18 (0.87,1.59) | 0.28 |
| Non-melanoma skin | 359 (11.5) | 249 (8.0) | 0.84 (0.62,1.15) | 0.28 | 1.05 (0.78,1.41) | 0.74 |
| Breast | 100 (3.2) | 69 (2.2) | 0.84 (0.55,1.29) | 0.42 | 0.90 (0.60,1.33) | 0.59 |
| Upper GI and hepatobiliary | 186 (6.0) | 181 (5.8) | 1.18 (0.84,1.66) | 0.34 | 1.12 (0.81,1.54) | 0.49 |
| Prostate | 142 (4.6) | 110 (3.5) | 0.94 (0.64,1.37) | 0.75 | 1.15 (0.81,1.64) | 0.44 |
| Lung | 43 (1.4) | 33 (1.1) | 0.93 (0.53,1.64) | 0.81 | 1.04 (0.61,1.75) | 0.89 |
| Melanoma | 29 (0.9) | 20 (0.6) | 0.84 (0.43,1.61) | 0.60 | 1.03 (0.54,1.94) | 0.92 |
| **Time from the First Cancer** |  |  |  |  |  |  |
| <6m | 446 (14.3) | 509 (16.3) | — |  |  |  |
| 6–12m | 158 (5.1) | 130 (4.2) | 0.72 (0.54,0.97) | **0.03** |  |  |
| 1–2y | 150 (4.8) | 146 (4.7) | 0.85 (0.63,1.15) | 0.29 |  |  |
| 2–5y | 303 (9.7) | 247 (7.9) | 0.72 (0.57,0.91) | **0.006** |  |  |
| >5y | 560 (17.9) | 471 (15.1) | 0.74 (0.61,0.9) | **0.003** |  |  |
| **Years since the First Cancer Diagnostic Code, per year** | 2.4 (0.4,7.5) | 1.8 (0.3,6.7) | 0.98 (0.97,1) | **0.03** | 1.02 (1.00,1.04) | **0.05** |
| **Active Chemotherapy Days in the Last Year, per 30 days** | 0 (0,65) | 0 (0,95) | 1.49 (1.11,2.01) | **0.008** |  |  |
| **Active Chemotherapy in Last Year** | 524 (16.8) | 618 (19.8) | 1.47 (1.24,1.74) | **<0.001** |  |  |
| **1-year-look-back Charlson Score, per 1 score** | 2 (1, 3) | 2 (1, 3) | 1.04 (0.98,1.09) | 0.21 |  |  |
| **IMD Percentile, per 1 Percentile (17 unknown)** | 78.0 (59.7,89.9) | 77.5 (59.5,89.6) | 1 (1, 1) | 0.52 |  |  |
| **Healthcare & Clinical History:** |  |  |  |  |  |  |
| **Number of Positive Urine Cultures with Enterobacterales in Last Year, per 1 culture** | 0 (0, 0) | 0 (0, 0) | 1.09 (0.96,1.23) | 0.19 |  |  |
| **Positive CSU Cultures in Last Year** | 120 (3.8) | 112 (3.6) | 0.99 (0.74,1.33) | 0.96 |  |  |
| **1-year-look-back Frailty Score, per 10 scores** | 9.4 (3.3,17.2) | 9.5 (4.1,17.3) | 1.02 (0.95,1.1) | 0.58 |  |  |
| **Days in Hospital before Collection Date in Last Year, per 10 days** | 6 (0,18) | 12 (1,30) | 1.13 (1.09,1.17) | **<0.001** |  |  |
| **Days in ICU before Collection Date in Last Year, per 1 day** | 0 (0, 0) | 0 (0, 0) | 1.08 (1.04,1.12) | **<0.001** |  |  |
| **Blood Infection Onset** |  |  |  |  |  |  |
| Within 48h of Current Admission (Community) | 1,283 (41.1) | 986 (31.6) | — |  | — |  |
| Beyond 48h of Current Admission (Hospital) | 334 (10.7) | 517 (16.6) | 2 (1.68,2.38) | **<0.001** | 1.49 (1.23,1.82) | **<0.001** |
| **Antibiotics Exposure:** |  |  |  |  |  |  |
| **Days on Non-Co-amoxiclav Antibiotics in Last Year, per 10 days** | 0 (0,0) | 0 (0,2) | 1.32 (1.21,1.44) | **<0.001** |  |  |
| **Days on Co-amoxiclav in Last Year, per 1 day** | 0 (0,0) | 0 (0,0) | 1.08 (1.05,1.1) | **<0.001** | 1.05 (1.02,1.08) | **<0.001** |
| **Days Since Last Antibiotic Used in Last Year, per 30 days** | 365 (365,365) | 365 (91,365) | **Non-linear** | |  |  |
| **Previous Resistance:** |  |  |  |  |  |  |
| **Previous Resistance in the Last Year** |  |  |  |  |  |  |
| No Previous AST Result/Positive Isolates, from Enterobacterales in Any Cultures | 1,088 (34.9) | 919 (29.5) | — |  | — |  |
| Resistance to Co-amoxiclav in Enterobacterales in Any Cultures | 175 (5.6) | 429 (13.8) | 2.91 (2.36,3.59) | **<0.001** | 2.44 (1.95,3.05) | **<0.001** |
| Resistance Only to Non-Co-amoxiclav in Enterobacterales in Any Cultures³ | 237 (7.6) | 111 (3.6) | 0.55 (0.43,0.71) | **<0.001** | 0.52 (0.40,0.68) | **<0.001** |
| Susceptible to All Tested Antimicrobials in Enterobacterales in Any Cultures | 117 (3.8) | 44 (1.4) | 0.45 (0.3,0.67) | **<0.001** | 0.44 (0.30,0.66) | **<0.001** |
| **Microbiological Cultures Sent:** |  |  |  |  |  |  |
| **Number of Urine Samples Taken in Last Year, per 10 cultures** | 1 (0, 3) | 2 (1, 4) | 1.99 (1.51,2.63) | **<0.001** |  |  |
| **Number of Neutrophil Tests in Last Year, per 10 tests** | 12 (5, 23) | 17 (8, 33) | 1.2 (1.15,1.24) | **<0.001** |  |  |
| **Number of Blood Samples Taken in Last Year, per 1 sample Study** | 1 (0, 2) | 2 (0, 5) | 1.07 (1.05,1.09) | **<0.001** | 1.03 (1.01,1.05) | **0.003** |
| **Timeline:** |  |  |  |  |  |  |
| **Calendar Days of Collection Date Since 01/04/2015, per year** | 2,142 (1,080,2,938) | 1,858 (966,2,807) | 0.95 (0.93,0.98) | **<0.001** | 0.95 (0.92,0.97) | **<0.001** |
| ¹n (%); Median (Q1, Q3) | | | | | | |
| ²Abbreviations: CI = Confidence Interval, OR = Odds Ratio | | | | | | |
| ³Equivalent to fully susceptible to Co-amoxiclav and resistant to any non-Co-amoxiclav in this dataset. | | | | | | |

Table S3: Associations (odds ratios and 95% confidence intervals) between potential risk factors and Enterobacterales-Co-amoxiclav in univariable and multivariable backward selection models.

| Fluoroquinolone in Enterobacterales positive blood cultures (N=3,138) | | | | | | |
| --- | --- | --- | --- | --- | --- | --- |
|  | **Susceptible N = 2,759**¹ | **Resistant N = 379**¹ | **Univariable (unadjusted)** | | **Multivariable (adjusted) with Backward Selection** | |
| **Variable** |  |  | **OR (95% CI)**² | **p-value** | **OR (95% CI)**² | **p-value** |
| **Demographics Characteristics and Cancer Related:** |  |  |  |  |  |  |
| **Age Group at Bloodstream Infection** |  |  |  |  |  |  |
| 60-80 | 1,260 (40.2) | 160 (5.1) | — |  |  |  |
| <40 | 92 (2.9) | 28 (0.9) | 2.48 (1.43,4.28) | **0.001** |  |  |
| 40-60 | 421 (13.4) | 85 (2.7) | 1.58 (1.11,2.24) | **0.03** |  |  |
| 80+ | 986 (31.4) | 106 (3.4) | 0.87 (0.65,1.16) | 0.46 |  |  |
| **Age at Bloodstream Infection, per 10 years** | 75.1 (64.0,83.9) | 71.7 (57.5,81.2) | 0.84 (0.78,0.91) | **<0.001** | 0.86 (0.77,0.96) | **0.005** |
| **Sex** |  |  |  |  |  |  |
| M | 1,689 (53.8) | 236 (7.5) | — |  | — |  |
| F | 1,070 (34.1) | 143 (4.6) | 0.95 (0.73,1.24) | 0.71 |  |  |
| **Ethnicity** |  |  |  |  |  |  |
| White | 2,214 (70.6) | 280 (8.9) | — |  | — |  |
| Non-white | 97 (3.1) | 32 (1.0) | 2.71 (1.71,4.31) | **<0.001** |  |  |
| Missing | 448 (14.3) | 67 (2.1) | 1.18 (0.84,1.67) | 0.34 |  |  |
| **Cancer Group** |  |  |  |  |  |  |
| Colorectal | 283 (9.0) | 30 (1.0) | — |  | — |  |
| Others | 492 (15.7) | 73 (2.3) | 1.32 (0.81,2.15) | 0.26 | 0.91 (0.54,1.54) | 0.73 |
| Lymphoid/Haematopoietic | 613 (19.5) | 116 (3.7) | 1.75 (1.11,2.77) | **0.02** | 1.48 (0.91,2.41) | 0.11 |
| Non-melanoma skin | 560 (17.8) | 50 (1.6) | 0.84 (0.49,1.43) | 0.52 | 0.91 (0.53,1.55) | 0.72 |
| Breast | 155 (4.9) | 16 (0.5) | 0.97 (0.51,1.85) | 0.93 | 0.86 (0.43,1.70) | 0.66 |
| Upper GI and hepatobiliary | 318 (10.1) | 52 (1.7) | 1.55 (0.9,2.66) | 0.11 | 1.54 (0.91, 2.61) | 0.11 |
| Prostate | 219 (7.0) | 35 (1.1) | 1.5 (0.86,2.62) | 0.15 | 1.41 (0.79,2.54) | 0.25 |
| Lung | 73 (2.3) | 4 (0.1) | 0.52 (0.17,1.53) | 0.23 | 0.52 (0.18,1.47) | 0.22 |
| Melanoma | 46 (1.5) | 3 (0.1) | 0.61 (0.18,2.12) | 0.44 | 0.83 (0.24,2.93) | 0.77 |
| **Time from the First Cancer** |  |  |  |  |  |  |
| <6m | 816 (26.0) | 148 (4.7) | — |  |  |  |
| 6–12m | 265 (8.4) | 24 (0.8) | 0.51 (0.32,0.83) | **0.007** |  |  |
| 1–2y | 258 (8.2) | 39 (1.2) | 0.81 (0.51,1.29) | 0.37 |  |  |
| 2–5y | 486 (15.5) | 65 (2.1) | 0.75 (0.52,1.09) | 0.13 |  |  |
| >5y | 934 (29.8) | 103 (3.3) | 0.62 (0.46,0.84) | **0.002** |  |  |
| **Years since the First Cancer Diagnostic Code, per year** | 2.2 (0.3,7.2) | 1.5 (0.2,5.9) | 0.98 (0.95,1) | 0.07 |  |  |
| **Active Chemotherapy Days in the Last Year, per 30 days** | 0 (0,81) | 0 (0,8) | 1 (0.96,1.04) | 0.92 |  |  |
| **Active Chemotherapy in Last Year** | 1,012 (32.2) | 137 (4.4) | 0.97 (0.75, 1.26) | 0.83 | 0.55 (0.39, 0.77) | **<0.001** |
| **1-year-look-back Charlson Score, per 1 score** | 2 (1,3) | 2 (1,3) | 1.05 (0.96,1.15) | 0.32 |  |  |
| **IMD Percentile, per 1 percentile (17 unknown)** | 78.0 (59.6,89.8) | 76.0 (59.6,87.5) | 1 (1,1.01) | 0.91 |  |  |
| **Healthcare & Clinical History:** |  |  |  |  |  |  |
| **Number of Positive Urine Cultures with Enterobacterales in Last Year, per 1 culture** | 0 (0,0) | 0 (0,1) | 1.49 (1.26,1.75) | **<0.001** |  |  |
| **Positive CSU Cultures in Last Year** | 199 (6.3) | 34 (1.1) | 1.29 (0.85,1.98) | 0.23 |  |  |
| **1-year-look-back Frailty Score, per 10 scores** | 9.3 (3.6,17.3) | 9.8 (3.2,16.7) | 0.94 (0.83,1.06) | 0.31 |  |  |
| **Days in Hospital before Collection Date in Last Year, per 10 days** | 8 (0, 23) | 12 (0, 33) | 1.09 (1.04,1.14) | **<0.001** |  |  |
| **Days in ICU before Collection Date in Last Year, per 1 day** | 0 (0,0) | 0 (0,0) | 1.05 (1,1.1) | **0.05** |  |  |
| **Blood Infection Onset** |  |  |  |  |  |  |
| Within 48h of Current Admission (Community) | 2,012 (64.1) | 267 (8.5) | — |  | — |  |
| Beyond 48h of Current Admission (Hospital) | 747 (23.8) | 112 (3.6) | 1.17 (0.9,1.53) | 0.24 |  |  |
| **Antibiotics Exposure:** |  |  |  |  |  |  |
| **Days on Non-Fluoroquinolone Antibiotics in Last Year, per 10 days** | 0 (0, 0) | 0 (0, 2) | 1.2 (1.1,1.31) | **<0.001** |  |  |
| **Days on Fluoroquinolone in Last Year, per 1 day** | 0 (0, 0) | 0 (0, 0) | 1.25 (1.12,1.4) | **<0.001** | 1.26 (1.11,1.44) | **<0.001** |
| **Days Since Last Antibiotic Used in Last Year, per 30 days** | 365 (365,365) | 365 (101,365) | 0.98 (0.95,1) | 0.06 |  |  |
| **Previous Resistance:** |  |  |  |  |  |  |
| **Previous Resistance in the Last Year** |  |  |  |  |  |  |
| No Previous AST Result/Positive Isolates, from Enterobacterales in Any Cultures | 1,818 (57.9) | 202 (6.4) | — |  | — |  |
| Resistance to Fluoroquinolone in Enterobacterales in Any Cultures | 59 (1.9) | 124 (4.0) | 18.79 (12.46,28.34) | **<0.001** | 15.23 (9.82,23.62) | **<0.001** |
| Resistance Only to Non-Fluoroquinolone in Enterobacterales in Any Cultures³ | 728 (23.2) | 45 (1.4) | 0.57 (0.4,0.81) | **0.002** | 0.45 (0.31,0.67) | **<0.001** |
| Susceptible to All Tested Antimicrobials in Enterobacterales in Any Cultures | 154 (4.9) | 8 (0.3) | 0.48 (0.23,0.99) | **0.05** | 0.47 (0.23,0.97) | **0.005** |
| **Microbiological Cultures Sent:** |  |  |  |  |  |  |
| **Number of Urine Samples Taken in Last Year, per 10 cultures** | 1 (0,3) | 2 (1,5) | 2.88 (2.02,4.1) | **<0.001** | 1.93 (1.14,3.25) | **0.01** |
| **Number of Neutrophil Tests in Last Year, per 10 tests** | 13 (6,27) | 16 (7,34) | 1.12 (1.06,1.18) | **<0.001** |  |  |
| **Number of Blood Samples Taken in Last Year, per 1 sample Study** | 1 (0,3) | 2 (0,6) | 1.03 (1.02,1.05) | **<0.001** |  |  |
| **Timeline:** |  |  |  |  |  |  |
| **Calendar Days of Collection Date Since 01/04/2015, per year** | 2,032 (1,037,2,897) | 1,771 (978,2,806) | 0.96 (0.92,1.03) | 0.07 | 0.94 (0.90,0.98) | **0.01** |
| ¹n (%); Median (Q1, Q3) | | | | | | |
| ²Abbreviations: CI = Confidence Interval, OR = Odds Ratio | | | | | | |
| ³Equivalent to fully susceptible to Fluoroquinolone and resistant to any non-Fluoroquinolone in this dataset. | | | | | | |

Table S4: Associations (odds ratios and 95% confidence intervals) between potential risk factors and Enterobacterales-Fluoroquinolone bacteraemia in univariable and multivariable backward selection models.

| Trimethoprim/Sulfamethoxazole in Enterobacterales positive blood cultures (N=3,108) | | | | | | |
| --- | --- | --- | --- | --- | --- | --- |
|  | **Susceptible N = 2,407**¹ | **Resistant N = 701**¹ | **Univariable (unadjusted)** | | **Multivariable (adjusted) with Backward Selection** | |
| **Variable** |  |  | **OR (95% CI)**² | **p-value** | **OR (95% CI)**² | **p-value** |
| **Demographics Characteristics and Cancer Related:** |  |  |  |  |  |  |
| **Age Group at Bloodstream Infection** |  |  |  |  |  |  |
| 60-80 | 1,106 (35.6) | 298 (9.6) | — |  |  |  |
| <40 | 76 (2.4) | 42 (1.4) | 2.08 (1.29,3.35) | **0.003** |  |  |
| 40-60 | 366 (11.8) | 135 (4.3) | 1.35 (1.01,1.8) | **0.04** |  |  |
| 80+ | 859 (27.6) | 226 (7.3) | 0.98 (0.78,1.22) | 0.84 |  |  |
| **Age at Bloodstream Infection, per 10 years** | 75.1 (64.0,83.9) | 72.4 (60.0,82.0) | 0.89 (0.83,0.95) | **<0.001** |  |  |
| **Sex** |  |  |  |  |  |  |
| M | 1,497 (48.2) | 408 (13.1) | — |  | — |  |
| F | 910 (29.3) | 293 (9.4) | 1.19 (0.97,1.46) | 0.09 |  |  |
| **Ethnicity** |  |  |  |  |  |  |
| White | 1,941 (62.5) | 531 (17.1) | — |  | — |  |
| Non-white | 84 (2.7) | 44 (1.4) | 1.97 (1.27,3.06) | **0.003** |  |  |
| Missing | 382 (12.3) | 126 (4.1) | 1.21 (0.92,1.58) | 0.16 |  |  |
| **Cancer Group** |  |  |  |  |  |  |
| Colorectal | 256 (8.2) | 52 (1.7) | — |  | — |  |
| Others | 427 (13.7) | 134 (4.3) | 1.52 (1.04,2.23) | **0.03** | 1.15 (0.75,1.75) | 0.52 |
| Lymphoid/Haematopoietic | 487 (15.7) | 234 (7.5) | 2.36 (1.64,3.39) | **<0.001** | 2.07 (1.40, .06) | **<0.001** |
| Non-melanoma skin | 497 (16.0) | 109 (3.5) | 1.08 (0.73, .58) | 0.71 | 1.06 (0.70,1.61) | 0.78 |
| Breast | 127 (4.1) | 41 (1.3) | 1.58 (0.94,2.68) | 0.09 | 1.29 (0.74,2.23) | 0.37 |
| Upper GI and hepatobiliary | 308 (9.9) | 60 (1.9) | 0.96 (0.61,1.52) | 0.87 | 0.94 (0.59,1.50) | 0.79 |
| Prostate | 205 (6.6) | 46 (1.5) | 1.1 (0.69,1.75) | 0.69 | 1.03 (0.63,1.66) | 0.91 |
| Lung | 60 (1.9) | 16 (0.5) | 1.31 (0.65,2.62) | 0.45 | 1.31 (0.66,2.60) | 0.44 |
| Melanoma | 40 (1.3) | 9 (0.3) | 1.1 (0.48,2.52) | 0.82 | 1.15 (0.50,2.64) | 0.74 |
| **Time from the First Cancer** |  |  |  |  |  |  |
| <6m | 738 (23.7) | 213 (6.9) | — |  |  |  |
| 6–12m | 224 (7.2) | 66 (2.1) | 1.04 (0.72,1.49) | 0.84 |  |  |
| 1–2y | 234 (7.5) | 63 (2.0) | 0.95 (0.67,1.36) | 0.79 |  |  |
| 2–5y | 405 (13.0) | 141 (4.5) | 1.22 (0.92,1.61) | 0.16 |  |  |
| >5y | 806 (25.9) | 218 (7.0) | 0.95 (0.74,1.21) | 0.67 |  |  |
| **Years since the First Cancer Diagnostic Code, per year** | 2.0 (0.3,6.9) | 2.1 (0.3,6.9) | 1 (0.98,1.02) | 0.77 |  |  |
| **Active Chemotherapy in Last Year** | 854 (27.5) | 285 (9.2) | 1.24 (1.01,1.51) | **0.04** |  |  |
| **Active Chemotherapy Days in the Last Year, per 30 days** | 0 (0, 69) | 0 (0, 108) | 1.05 (1.02,1.08) | **0.001** |  |  |
| **1-year-look-back Charlson Score, per 1 score** | 2 (1,3) | 2 (1,3) | 1 (0.93,1.07) | 0.95 |  |  |
| **IMD Percentile, per 1 percentile (16 unknown)** | 77.7 (59.5,89.8) | 77.7 (60.7,89.8) | 1 (1,1.01) | 0.42 |  |  |
| **Healthcare & Clinical History:** |  |  |  |  |  |  |
| **Number of Positive Urine Cultures with Enterobacterales in Last Year, per 1 culture** | 0 (0,0) | 0 (0,1) | 1.45 (1.26,1.67) | **<0.001** |  |  |
| **Positive CSU Cultures in Last Year** | 167 (5.4) | 65 (2.1) | 1.39 (0.98,1.97) | 0.06 |  |  |
| **1-year-look-back Frailty Score, per 10 scores** | 9.1 (3.5,16.6) | 9.7 (3.6,17.7) | 1.05 (0.96,1.16) | 0.28 |  |  |
| **Days in Hospital before Collection Date in Last Year, per 10 days** | 7 (0,22) | 10 (0,27) | 1.06 (1.02,1.1) | **0.002** |  |  |
| **Days in ICU before Collection Date in Last Year, per 1 day** | 0 (0,0) | 0 (0,0) | 1.02 (0.98,1.06) | 0.42 |  |  |
| **Blood Infection Onset** |  |  |  |  |  |  |
| Within 48h of Current Admission (Community) | 1,749 (56.3) | 506 (16.3) | — |  | — |  |
| Beyond 48h of Current Admission (Hospital) | 658 (21.2) | 195 (6.3) | 1.04 (0.85,1.29) | 0.67 |  |  |
| **Antibiotics Exposure:** |  |  |  |  |  |  |
| **Days on Non-Trimethoprim/Sulfamethoxazole Antibiotics in Last Year, per 10 days** | 0 (0,0) | 0 (0,0) | 1.09 (1.01,1.18) | **0.04** | 0.89 (0.79,0.99) | **0.04** |
| **Days on Trimethoprim/Sulfamethoxazole in Last Year, per 1 day** | 0 (0,0) | 0 (0,0) | 1.14 (1.08,1.21) | **<0.001** | 1.15 (1.07,1.24) | **<0.001** |
| **Days Since Last Antibiotic Used in Last Year, per 30 days** | 365 (365,365) | 365 (365,365) | 0.99 (0.97,1.01) | 0.47 |  |  |
| **Previous Resistance:** |  |  |  |  |  |  |
| **Previous Resistance in the Last Year** |  |  |  |  |  |  |
| No Previous AST Result/Positive Isolates, from Enterobacterales in Any Cultures | 1,599 (51.4) | 401 (12.9) | — |  | — |  |
| Resistance to Trimethoprim/Sulfamethoxazole in Enterobacterales in Any Cultures | 80 (2.6) | 212 (6.8) | 10.43 (7.58,4.34) | **<0.001** | 8.84 (6.34, 2.33) | **<0.001** |
| Resistance Only to Non-Trimethoprim/Sulfamethoxazole in Enterobacterales in Any Cultures³ | 584 (18.8) | 73 (2.3) | 0.5 (0.38, 0.67) | **<0.001** | 0.44 (0.32,0.60) | **<0.001** |
| Susceptible to All Tested Antimicrobials in Enterobacterales in Any Cultures | 144 (4.6) | 15 (0.5) | 0.42 (0.23,0.74) | **0.003** | 0.40 (0.22,0.72) | **0.002** |
| **Microbiological Cultures Sent:** |  |  |  |  |  |  |
| **Number of Neutrophil Tests in Last Year, per 10 tests** | 13 (6,26) | 16 (7,33) | 1.11 (1.07,1.16) | **<0.001** |  |  |
| **Number of Blood Samples Taken in Last Year, per 1 sample** | 1 (0,3) | 1 (0,4) | 1.03 (1.01,1.04) | **<0.001** |  |  |
| **Number of Urine Samples Taken in Last Year, per 10 cultures** | 1 (0,3) | 2 (1,5) | 2.56 (1.88,3.49) | **<0.001** | 1.91 (1.26,2.89) | **0.002** |
| **Study Timeline:** |  |  |  |  |  |  |
| **Calendar Days of Collection Date Since 01/04/2015, per year** | 1,968 (982,2,774) | 1,661 (920,2,584) | **Non-linear** | | 0.93 (0.90,0.97) | **<0.001** |
| ¹n (%); Median (Q1, Q3) | | | | | | |
| ²Abbreviations: CI = Confidence Interval, OR = Odds Ratio | | | | | | |
| ³Equivalent to fully susceptible to Trimethoprim/Sulfamethoxazole and resistant to any non-Trimethoprim/Sulfamethoxazole in this dataset. | | | | | | |

Table S5: Associations (odds ratios and 95% confidence intervals) between potential risk factors and Enterobacterales-Trimethoprim/Sulfamethoxazole in univariable and multivariable backward selection models.

| Third-generation cephalosporin in Enterobacterales positive blood cultures (N=3,139) | | | | | | |
| --- | --- | --- | --- | --- | --- | --- |
|  | **Susceptible N = 2,716**¹ | **Resistant N = 423**¹ | **Univariable (unadjusted)** | | **Multivariable (adjusted) with Backward Selection** | |
| **Variable** |  |  | **OR (95% CI)**² | **p-value** | **OR (95% CI)**² | **p-value** |
| **Demographics Characteristics and Cancer Related:** |  |  |  |  |  |  |
| **Age Group at Bloodstream Infection** |  |  |  |  |  |  |
| 60-80 | 1,223 (39.0) | 197 (6.3) | — |  |  |  |
| <40 | 89 (2.8) | 31 (1.0) | 2.16 (1.24,3.76) | **0.006** |  |  |
| 40-60 | 417 (13.3) | 91 (2.9) | 1.35 (0.97,1.89) | 0.07 |  |  |
| 80+ | 987 (31.4) | 104 (3.3) | 0.65 (0.49,0.87) | **0.004** |  |  |
| **Age at Bloodstream Infection, per 10 years** | 75.2 (64.0,84.1) | 70.3 (58.2,79.8) | 0.82 (0.76,0.88) | **<0.001** | 0.91 (0.83,0.99) | **0.05** |
| **Sex** |  |  |  |  |  |  |
| M | 1,652 (52.6) | 274 (8.7) | — |  | — |  |
| F | 1,064 (33.9) | 149 (4.7) | 0.86 (0.67,1.11) | 0.26 |  |  |
| **Ethnicity** |  |  |  |  |  |  |
| White | 2,181 (69.5) | 312 (9.9) | — |  | — |  |
| Non-white | 98 (3.1) | 32 (1.0) | 2.34 (1.45,3.77) | **<0.001** |  |  |
| Missing | 437 (13.9) | 79 (2.5) | 1.23 (0.89,1.72) | 0.21 |  |  |
| **Cancer Group** |  |  |  |  |  |  |
| Colorectal | 272 (8.7) | 40 (1.3) | — |  | — |  |
| Others | 476 (15.2) | 89 (2.8) | 1.27 (0.81,2) | 0.297 | 1.20 (0.75,1.94) | 0.45 |
| Lymphoid/Haematopoietic | 610 (19.4) | 121 (3.9) | 1.35 (0.88,2.06) | 0.17 | 0.97 (0.60,1.57) | 0.89 |
| Non-melanoma skin | 553 (17.6) | 57 (1.8) | 0.72 (0.45,1.15) | 0.17 | 0.85 (0.51,1.39) | 0.51 |
| Breast | 158 (5.0) | 13 (0.4) | 0.56 (0.29,1.08) | 0.09 | 0.61 (0.30,1.25) | 0.18 |
| Upper GI and hepatobiliary | 309 (9.8) | 61 (1.9) | 1.39 (0.84,2.28) | 0.21 | 1.48 (0.88,2.49) | 0.14 |
| Prostate | 222 (7.1) | 32 (1.0) | 1.01 (0.57,1.76) | 0.99 | 1.23 (0.69,2.21) | 0.48 |
| Lung | 73 (2.3) | 4 (0.1) | 0.38 (0.11,1.38) | 0.14 | 0.34 (0.09,1.31) | 0.12 |
| Melanoma | 43 (1.4) | 6 (0.2) | 0.97 (0.39,2.4) | 0.95 | 1.28 (0.45,3.11) | 0.62 |
| **Time from the First Cancer** |  |  |  |  |  |  |
| <6m | 809 (25.8) | 156 (5.0) | — |  |  |  |
| 6–12m | 253 (8.1) | 37 (1.2) | 0.76 (0.48,1.19) | 0.23 |  |  |
| 1–2y | 250 (8.0) | 47 (1.5) | 0.97 (0.63,1.52) | 0.91 |  |  |
| 2–5y | 476 (15.2) | 75 (2.4) | 0.82 (0.58,1.15) | 0.25 |  |  |
| >5y | 928 (29.6) | 108 (3.4) | 0.61 (0.45,0.83) | **0.002** |  |  |
| **Time since the First Cancer Diagnostic Code, per year** | 2.2 (0.3,7.2) | 1.3(0.3,5.2) | **Non-linear** | |  |  |
| **Active Chemotherapy Days in the Last Year, per 30 days** | 0 (0, 77) | 0 (0, 108) | 1.05 (1.01,1.08) | **0.009** |  |  |
| **Active Chemotherapy in Last Year** | 971 (30.9) | 180 (5.7) | 1.33 (1.04,1.7) | **0.02** |  |  |
| **1-year-look-back Charlson Score, per 1 score** | 2 (1, 3) | 2 (1,3) | 1.08 (0.99,1.17) | 0.07 |  |  |
| **IMD Percentile, per 1 Percentile (17 unknown)** | 78.0 (60.0,90.0) | 77.0 (61.0,88.0) | 1 (0.99,1) | 0.73 |  |  |
| **Healthcare & Clinical History:** |  |  |  |  |  |  |
| **Number of Positive Urine Cultures in Enterobacterales in Last Year, per 1 culture** | 0 (0, 0) | 0 (0,1) | 1.33 (1.12,1.59) | **0.001** |  |  |
| **Positive CSU Cultures in Last Year** | 198 (6.3) | 35 (1.1) | 1.15 (0.77,1.71) | 0.50 |  |  |
| **1-year-look-back Frailty Score, per 10 scores** | 9.1 (3.5,16.8) | 11.5 (4.3,18.0) | 1.11 (1, 1.23) | 0.06 |  |  |
| **Days in Hospital before Collection Date in Last Year, per 10 days** | 7 (0,2) | 16 (1,41) | 1.13 (1.09,1.18) | **<0.001** |  |  |
| **Days in ICU before Collection Date in Last Year, per 1 day** | 0 (0,0) | 0 (0,0) | 1.08 (1.04,1.12) | **<0.001** |  |  |
| **Blood Infection Onset** |  |  |  |  |  |  |
| Within 48h of Current Admission (Community) | 2,003 (63.8) | 277 (8.8) | — |  | — |  |
| Beyond 48h of Current Admission (Hospital) | 713 (22.7) | 146 (4.7) | 1.48 (1.16,1.88) | **0.001** |  |  |
| **Antibiotics Exposure:** |  |  |  |  |  |  |
| **Days on Any Non-Third-generation Cephalosporin in Last Year, per 10 days** | 0 (0, 0) | 0 (0,7) | 1.26 (1.16,1.38) | **<0.001** |  |  |
| **Days on Third-generation Cephalosporin in Last Year, per 1 day** | 0 (0, 0) | 0 (0,0) | 1.26 (1.13,1.4) | **<0.001** | 1.21 (1.09, 1.35) | **<0.001** |
| **Days Since Last Antibiotic Used in Last Year, per 30 days** | 365 (365,365) | 365 (47,365) | 0.96 (0.94,0.98) | **0.001** |  |  |
| **Previous Resistance:** |  |  |  |  |  |  |
| **Previous Resistance in the Last Year** |  |  |  |  |  |  |
| No Previous AST Result/Positive Isolates, from Enterobacterales in Any Cultures | 1,802 (57.4) | 217 (6.9) | — |  | — |  |
| Resistance to Third-generation Cephalosporin in Enterobacterales in Any Cultures | 83 (2.6) | 131 (4.2) | 12.97 (9.11,18.48) | **<0.001** | 10.61 (7.31,15.41) | **<0.001** |
| Resistance Only to Non-Third-generation Cephalosporin in Enterobacterales in Any Cultures³ | 683 (21.8) | 61 (1.9) | 0.73 (0.54,1) | **0.05** | 0.62 (0.45,0.87) | **0.005** |
| Susceptible to All Tested Antimicrobials in Enterobacterales in Any Cultures⁴ | 148 (4.7) | 14 (0.4) | 0.79 (0.39,1.57) | 0.49 | 0.77 (0.38,1.56) | 0.46 |
| **Microbiological Cultures Sent:** |  |  |  |  |  |  |
| **Number of Urine Samples Taken in Last Year, per 10 cultures** | 1 (0,3) | 3 (1,5) | 3.19 (2.23,4.58) | **<0.001** |  |  |
| **Number of Neutrophil Tests in Last Year, per 10 tests** | 13 (6,26) | 21 (10,42) | 1.19 (1.14,1.25) | **<0.001** |  |  |
| **Number of Blood Samples Taken in Last Year, per 1 sample Study** | 1 (0,3) | 2 (0,7) | 1.06 (1.04,1.07) | **<0.001** | 1.03 (1.01,1.05) | **0.003** |
| **Timeline:** |  |  |  |  |  |  |
| **Calendar Days of Collection Date Since 01/04/2015, per year** | 2,029 (1,022,2,889) | 1,942 (1,102,2,829) | 0.99 (0.95,1.04) | 0.81 |  |  |
| ¹n (%); Median (Q1, Q3) | | | | | | |
| ²Abbreviations: CI = Confidence Interval, OR = Odds Ratio | | | | | | |
| ³Equivalent to fully susceptible to Third-generation cephalosporin and resistant to any non-Third-generation cephalosporin in this dataset. ⁴2 had no previous isolates susceptible to 3GC. | | | | | | |

Table S6: Associations (odds ratios and 95% confidence intervals) between potential risk factors and Enterobacterales-Third-generation cephalosporin in univariable and multivariable backward selection models.

| Gentamicin in Enterobacterales positive blood cultures (N=3,138) | | | | | | |
| --- | --- | --- | --- | --- | --- | --- |
|  | **Susceptible N = 2,850**¹ | **Resistant N = 288**¹ | **Univariable (unadjusted)** | | **Multivariable (adjusted) with Backward Selection** | |
| **Variable** |  |  | **OR (95% CI)**² | **p-value** | **OR (95% CI)**² | **p-value** |
| **Demographics Characteristics and Cancer Related:** |  |  |  |  |  |  |
| **Age Group at Bloodstream Infection** |  |  |  |  |  |  |
| 60-80 | 1,280 (40.8) | 139 (4.4) | — |  |  |  |
| <40 | 105 (3.3) | 15 (0.5) | 1.21 (0.66,2.23) | 0.54 |  |  |
| 40-60 | 466 (14.9) | 42 (1.3) | 0.87 (0.56,1.36) | 0.55 |  |  |
| 80+ | 999 (31.8) | 92 (2.9) | 0.86 (0.63,1.17) | 0.34 |  |  |
| **Age at Bloodstream Infection, per 10 years** | 74.8 (63.4,83.6) | 73.6 (63.6,82.7) | 0.95 (0.87,1.04) | 0.24 |  |  |
| **Sex** |  |  |  |  |  |  |
| M | 1,735 (55.3) | 191 (6.1) | — |  | — |  |
| F | 1,115 (35.5) | 97 (3.1) | 0.79 (0.58,1.08) | 0.14 |  |  |
| **Ethnicity** |  |  |  |  |  |  |
| White | 2,262 (72.1) | 231 (7.4) | — |  | — |  |
| Non-white | 112 (3.6) | 18 (0.6) | 1.6 (0.79,3.22) | 0.19 |  |  |
| Missing | 476 (15.2) | 39 (1.2) | 0.81 (0.53,1.26) | 0.35 |  |  |
| **Cancer Group** |  |  |  |  |  |  |
| Colorectal | 292 (9.3) | 21 (0.7) | — |  | — |  |
| Others | 499 (15.9) | 66 (2.1) | 1.85 (1.06,3.24) | **0.03** | 1.45 (0.81,2.59) | 0.21 |
| Lymphoid/Haematopoietic | 648 (20.7) | 83 (2.6) | 1.8 (1.04,3.1) | **0.03** | 1.56 (0.87,2.77) | 0.13 |
| Non-melanoma skin | 566 (18.0) | 43 (1.4) | 1.05 (0.59,1.88) | 0.86 | 0.90 (0.47,1.69) | 0.74 |
| Breast | 161 (5.1) | 10 (0.3) | 0.86 (0.37,1.99) | 0.73 | 0.60 (0.22,1.66) | 0.33 |
| Upper GI and hepatobiliary | 347 (11.1) | 23 (0.7) | 0.92 (0.43,1.97) | 0.84 | 0.95 (0.46,1.97) | 0.89 |
| Prostate | 220 (7.0) | 33 (1.1) | 2.08 (1.12,3.86) | **0.02** | 1.71 (0.90,3.23) | 0.09 |
| Lung | 70 (2.2) | 7 (0.2) | 1.39 (0.51,3.74) | 0.52 | 1.28 (0.46,3.58) | 0.64 |
| Melanoma | 47 (1.5) | 2 (0.1) | 0.59 (0.14,2.56) | 0.48 | 0.65 (0.14,2.92) | 0.57 |
| **Time from the First Cancer** |  |  |  |  |  |  |
| <6m | 869 (27.7) | 96 (3.1) | — |  |  |  |
| 6–12m | 266 (8.5) | 23 (0.7) | 0.77 (0.46,1.29) | 0.32 |  |  |
| 1–2y | 265 (8.4) | 32 (1.0) | 1.01 (0.64,1.6) | 0.95 |  |  |
| 2–5y | 501 (16.0) | 50 (1.6) | 0.89 (0.6,1.34) | 0.59 |  |  |
| >5y | 949 (30.2) | 87 (2.8) | 0.81 (0.57,1.16) | 0.25 |  |  |
| **Years since the First Cancer Diagnostic Code, per year** | 2.1 (0.3,7.2) | 1.8 (0.3,6.6) | 0.99 (0.96,1.02) | 0.56 |  |  |
| **Active Chemotherapy Days in the Last Year, per 30 days** | 0 (0,81) | 0 (0,81.5) | 0.98 (0.94,1.03) | 0.45 |  |  |
| **Active Chemotherapy in Last Year** | 1,047 (33.4) | 104 (3.3) | 0.97 (0.73,1.31) | 0.86 |  |  |
| **1-year-look-back Charlson Score, per 1 score** | 2 (1,3) | 2 (1,3) | 1.05 (0.95,1.15) | 0.35 |  |  |
| **1-year-look-back Frailty Score, per 10 scores** | 9.0 (3.5,16.7) | 13.4 (5.5,22.7) | 1.39 (1.23,1.58) | **<0.001** | 1.30 (1.12, 1.50) | **<0.001** |
| **IMD Percentile, per 1 percentile (17 unknown)** | 77.7 (59.5,89.6) | 78.9 (62.2,91.4) | 1 (1,1.01) | 0.45 |  |  |
| **Healthcare & Clinical History:** |  |  |  |  |  |  |
| **Number of Positive Urine Cultures with Enterobacterales in Last Year, per 1 culture** | 0 (0,0) | 0 (0,1) | 1.44 (1.25,1.66) | **<0.001** |  |  |
| **Positive CSU Cultures in Last Year** | 189 (6.0) | 44 (1.4) | 2.55 (1.67,3.88) | **<0.001** |  |  |
| **Days in Hospital before Collection Date in Last Year, per 10 days** | 8 (0,22) | 15 (1,37) | 1.11 (1.06,1.16) | **<0.001** |  |  |
| **Days in ICU before Collection Date in Last Year, per 1 day** | 0 (0,0) | 0 (0,0) | 1.02 (0.97,1.07) | 0.36 |  |  |
| **Blood Infection Onset** |  |  |  |  |  |  |
| Within 48h of Current Admission (Community) | 2,066 (65.8) | 213 (6.8) | — |  | — |  |
| Beyond 48h of Current Admission (Hospital) | 784 (25.0) | 75 (2.4) | 0.93 (0.7,1.25) | 0.65 |  |  |
| **Antibiotics Exposure:** |  |  |  |  |  |  |
| **Days on Non-Gentamicin Antibiotics in Last Year, per 10 days** | 0 (0,0) | 0 (0,0) | 1.12 (1.01,1.24) | **0.04** |  |  |
| **Days on Gentamicin in Last Year, per 1 day** | 0 (0,0) | 0 (0,0) | 1.2 (1.01,1.43) | **0.04** |  |  |
| **Days Since Last Antibiotic Used in Last Year, per 30 days** | 365 (365,365) | 365 (365,365) | 0.99 (0.96,1.02) | 0.51 |  |  |
| **Previous Resistance:** |  |  |  |  |  |  |
| **Previous Resistance in the Last Year** |  |  |  |  |  |  |
| No Previous AST Result/Positive Isolates, from Enterobacterales in Any Cultures | 1,865 (59.4) | 153 (4.9) | — |  | — |  |
| Resistance to Gentamicin in Enterobacterales in Any Cultures | 74 (2.4) | 87 (2.8) | 14.27 (9.51,21.41) | **<0.001** | 11.17 (7.37,16.92) | **<0.001** |
| Resistance Only to Non-Gentamicin in Enterobacterales in Any Cultures³ | 760 (24.2) | 37 (1.2) | 0.6 (0.4,0.89) | **0.01** | 0.51 (0.34,0.79) | **0.002** |
| Susceptible to All Tested Antimicrobials in Enterobacterales in Any Cultures⁴ | 151 (4.8) | 11 (0.4) | 0.88 (0.46,1.68) | 0.71 | 0.79 (0.41,1.52) | 0.48 |
| **Microbiological Cultures Sent:** |  |  |  |  |  |  |
| **Number of Neutrophil Tests in Last Year, per 10 tests** | 13 (6,27) | 18 (8,39) | 1.13 (1.07,1.2) | **<0.001** |  |  |
| **Number of Blood Samples Taken in Last Year, per 1 sample** | 1 (0,3) | 2 (0,6) | 1.03 (1.01,1.05) | **0.002** |  |  |
| **Number of Urine Samples Taken in Last Year, per 10 cultures** | 1 (0,3) | 3 (1,6) | 3.62 (2.44,5.37) | **<0.001** |  |  |
| **Study Timeline:** |  |  |  |  |  |  |
| **Calendar Days of Collection Date Since 01/04/2015, per year** | 1,993 (1,019,2,866) | 2,204 (1,139,3,083) | 1.05 (1,1.1) | 0.06 |  |  |
| ¹n (%); Median (Q1, Q3) | | | | | | |
| ²Abbreviations: CI = Confidence Interval, OR = Odds Ratio | | | | | | |
| ³Equivalent to fully susceptible to Gentamicin and resistant to any non-Gentamicin in this dataset. ⁴2 had no previous isolates susceptible to Gentamicin. | | | | | | |

Table S7: Associations (odds ratios and 95% confidence intervals) between potential risk factors and Enterobacterales-Gentamicin in univariable and multivariable backward selection models.

| Piperacillin-tazobactam in Enterobacterales positive blood cultures (N=3,139) | | | | | | |
| --- | --- | --- | --- | --- | --- | --- |
|  | **Susceptible N = 2,775**¹ | **Resistant N = 364**¹ | **Univariable (unadjusted)** | | **Multivariable (adjusted) with Backward Selection** | |
| **Variable** |  |  | **OR (95% CI)**² | **p-value** | **OR (95% CI)**² | **p-value** |
| **Demographics Characteristics and Cancer Related:** |  |  |  |  |  |  |
| **Age Group at Bloodstream Infection** |  |  |  |  |  |  |
| 60-80 | 1,250 (39.8) | 170 (5.4) | — |  |  |  |
| <40 | 84 (2.7) | 36 (1.1) | 3.21 (2.06,4.99) | **<0.001** |  |  |
| 40-60 | 415 (13.2) | 92 (2.9) | 1.6 (1.17,2.18) | **0.003** |  |  |
| 80+ | 1,026 (32.7) | 66 (2.1) | 0.48 (0.34,0.66) | **<0.001** |  |  |
| **Age at Bloodstream Infection, per 10 years** | 75.4 (64.6,84.2) | 65.8 (53.1,77.4) | 0.71 (0.66,0.76) | **<0.001** | 0.83 (0.76,0.91) | **<0.001** |
| **Sex** |  |  |  |  |  |  |
| M | 1,716 (54.7) | 211 (6.7) | — |  | — |  |
| F | 1,059 (33.7) | 153 (4.9) | 1.15 (0.89,1.48) | 0.30 |  |  |
| **Ethnicity** |  |  |  |  |  |  |
| White | 2,227 (70.9) | 266 (8.5) | — |  | — |  |
| Non-white | 106 (3.4) | 24 (0.8) | 1.95 (1.21,3.15) | **0.006** |  |  |
| Missing | 442 (14.1) | 74 (2.4) | 1.4 (1,1.94) | **0.05** |  |  |
| **Cancer Group** |  |  |  |  |  |  |
| Colorectal | 277 (8.8) | 36 (1.1) | — |  | — |  |
| Others | 503 (16.0) | 61 (1.9) | 0.94 (0.58,1.51) | 0.796 | 0.95 (0.58,1.56) | 0.85 |
| Lymphoid/Haematopoietic | 585 (18.6) | 146 (4.7) | 1.86 (1.22,2.83) | **0.004** | 1.18 (0.75,1.86) | 0.48 |
| Non-melanoma skin | 567 (18.1) | 43 (1.4) | 0.58 (0.35,0.96) | **0.03** | 0.87 (0.52,1.45) | 0.59 |
| Breast | 158 (5.0) | 13 (0.4) | 0.63 (0.3,1.32) | 0.22 | 0.73 (0.34,1.54) | 0.41 |
| Upper GI and hepatobiliary | 323 (10.3) | 47 (1.5) | 1.12 (0.66,1.92) | 0.67 | 1.06 (0.61,1.83) | 0.84 |
| Prostate | 242 (7.7) | 12 (0.4) | 0.38 (0.19,0.78) | **0.008** | 0.55 (0.27,1.15) | 0.11 |
| Lung | 75 (2.4) | 2 (0.1) | 0.2 (0.03,1.53) | 0.12 | 0.22 (0.03,1.74) | 0.15 |
| Melanoma | 45 (1.4) | 4 (0.1) | 0.68 (0.23,2.04) | 0.49 | 0.92 (0.29,2.96) | 0.89 |
| **Time from the First Cancer** |  |  |  |  |  |  |
| <6m | 832 (26.5) | 133 (4.2) | — |  |  |  |
| 6–12m | 253 (8.1) | 36 (1.1) | 0.92 (0.6,1.41) | 0.69 |  |  |
| 1–2y | 248 (7.9) | 49 (1.6) | 1.28 (0.83,1.98) | 0.26 |  |  |
| 2–5y | 499 (15.9) | 52 (1.7) | 0.67 (0.46,0.97) | **0.03** |  |  |
| >5y | 943 (30.0) | 94 (3.0) | 0.64 (0.47,0.88) | **0.006** |  |  |
| **Years since the First Cancer Diagnostic Code, per year** | 2.2 (0.3,7.3) | 1.2 (0.2,5.2) | 0.97 (0.94,0.99) | **0.01** |  |  |
| **Active Chemotherapy Days in the Last Year, per 30 days** | 0 (0, 71) | 0 (0, 131) | 1.07 (1.03,1.1) | **<0.001** |  |  |
| **Active Chemotherapy in Last Year** | 973 (31.0) | 177 (5.6) | 1.76 (1.38,2.26) | **<0.001** |  |  |
| **1-year-look-back Charlson Score, per 1 score** | 2 (1, 3) | 2 (1, 3) | 1.04 (0.95,1.13) | 0.38 |  |  |
| **IMD Percentile, per 1 percentile (17 unknown)** | 78.0 (59.7,89.8) | 76.1 (56.2,88.5) | 1 (0.99,1) | 0.2 |  |  |
| **Healthcare & Clinical History:** |  |  |  | |  |  |
| **Number of Positive Urine Cultures with Enterobacterales in Last Year, per 1 culture** | 0 (0,0) | 0 (0,0) | 0.94 (0.78,1.13) | 0.49 |  |  |
| **Positive CSU Cultures in Last Year** | 209 (6.7) | 24 (0.8) | 0.97 (0.61,1.54) | 0.91 |  |  |
| **1-year-look-back Frailty Score, per 10 scores** | 9.3 (3.5,17.2) | 9.6 (4.0,17.5) | 0.97 (0.87,1.08) | 0.621 |  |  |
| **Days in Hospital before Collection Date in Last Year, per 10 days** | 7 (0,22) | 16 (3,40) | 1.14 (1.09,1.18) | **<0.001** |  |  |
| **Days in ICU before Collection Date in Last Year, per 1 day** | 0 (0,0) | 0 (0,0) | 1.09 (1.05,1.13) | **<0.001** |  |  |
| **Blood Infection Onset** |  |  |  |  |  |  |
| Within 48h of Current Admission (Community) | 2,100 (66.9) | 180 (5.7) | — |  | — |  |
| Beyond 48h of Current Admission (Hospital) | 675 (21.5) | 184 (5.9) | 3.14 (2.45,4) | **<0.001** | 1.93 (1.46,2.55) | **<0.001** |
| **Antibiotics Exposure:** |  |  |  | |  |  |
| **Days on Non-Piperacillin / Tazobactam Antibiotics in Last Year, per 10 days** | 0 (0,0) | 0 (0,8) | 1.27 (1.17,1.38) | **<0.001** |  |  |
| **Days on Piperacillin / Tazobactam in Last Year, per 1 day** | 0 (0,0) | 0 (0,0) | 1.08 (1.06,1.11) | **<0.001** |  |  |
| **Days Since Last Antibiotic Used in Last Year, per 30 days** | 365 (365,365) | 365 (9,365) | **Non-linear** | | 0.97 (0.95,0.99) | **0.01** |
| **Previous Resistance:** |  |  |  |  |  |  |
| **Previous Resistance in the Last Year** |  |  |  |  |  |  |
| No Previous AST Result/Positive Isolates, from Enterobacterales in Any Cultures | 1,815 (57.8) | 205 (6.5) | — |  | — |  |
| Resistance to Piperacillin/Tazobactam in Enterobacterales in Any Cultures | 100 (3.2) | 67 (2.1) | 5.94 (4.08,8.66) | **<0.001** | 4.75 (3.06,7.38) | **<0.001** |
| Resistance Only to Non-Piperacillin/Tazobactam in Enterobacterales in Any Cultures³ | 709 (22.6) | 81 (2.6) | 1.03 (0.77,1.39) | 0.84 | 1.05 (0.78,1.42) | 0.75 |
| Susceptible to All Tested Antimicrobials in Enterobacterales in Any Cultures⁴ | 151 (4.8) | 11 (0.4) | 0.65 (0.3,1.4) | 0.27 | 0.78 (0.37,1.63) | 0.51 |
| **Microbiological Cultures Sent:** |  |  |  |  |  |  |
| **Number of Neutrophil Tests in Last Year, per 10 tests** | 13 (6,26) | 22 (12,46) | 1.2 (1.15,1.26) | **<0.001** |  |  |
| **Number of Blood Samples Taken in Last Year, per 1 sample** | 1 (0,3) | 3 (1,9) | 1.06 (1.05,1.08) | **<0.001** |  |  |
| **Number of Urine Samples Taken in Last Year, per 10 cultures** | 1 (0,3) | 2 (1,4) | 2.29 (1.62,3.24) | **<0.001** |  |  |
| **Study Timeline:** |  |  |  |  |  |  |
| **Calendar Days of Collection Date Since 01/04/2015, per year** | 2,015 (1,054,2,897) | 1,977 (902,2,756) | 0.96 (0.93,1) | 0.06 |  |  |
| ¹n (%); Median (Q1, Q3) | | | | | | |
| ²Abbreviations: CI = Confidence Interval, OR = Odds Ratio | | | | | | |
| ³Equivalent to fully susceptible to Piperacillin-tazobactam and resistant to any non-Piperacillin-tazobactam in this dataset. | | | | | | |
| ⁴2 had no previous isolates susceptible to Piperacillin/Tazobactam. | | | | | | |

Table S8: Associations (odds ratios and 95% confidence intervals) between potential risk factors and Enterobacterales-Piperacillin-Tazobactam in univariable and multivariable backward selection models.

| Vancomycin in Enterococcus faecalis/faecium positive blood cultures (N=617) | | | | | | |
| --- | --- | --- | --- | --- | --- | --- |
|  | **Susceptible N = 488**¹ | **Resistant N = 129**¹ | **Univariable (unadjusted)** | | **Multivariable (adjusted) with Backward Selection** | |
| **Variable** |  |  | **OR (95% CI)**² | **p-value** | **OR (95% CI)**² | **p-value** |
| **Demographics Characteristics and Cancer Related:** |  |  |  |  |  |  |
| **Age Group at Bloodstream Infection** |  |  |  |  |  |  |
| 60-80 | 215 (34.8) | 49 (7.9) | — |  |  |  |
| <40 | 32 (5.2) | 31 (5.0) | 4.37 (2.28,8.38) | **<0.001** |  |  |
| 40-60 | 109 (17.7) | 40 (6.5) | 1.67 (1.02,2.73) | **0.04** |  |  |
| 80+ | 131 (21.4) | 9 (1.5) | 0.31 (0.14,0.71) | **0.005** |  |  |
| **Age at Bloodstream Infection, per 10 years** | 70.3 (58.4,80.5) | 58.8 (40.5,67.8) | 0.66 (0.59,0.74) | **<0.001** | 0.80 (0.69,0.93) | **0.002** |
| **Sex** |  |  |  |  |  |  |
| M | 309 (50.1) | 88 (14.3) | — |  | — |  |
| F | 179 (29.0) | 41 (6.6) | 0.78 (0.49,1.24) | 0.31 |  |  |
| **Ethnicity** |  |  |  |  |  |  |
| White | 367 (59.5) | 95 (15.4) | — |  | — |  |
| Non-white | 17 (2.8) | 7 (1.1) | 1.61 (0.63,4.1) | 0.32 |  |  |
| Missing | 104 (16.9) | 27 (4.4) | 1.04 (0.6,1.78) | 0.89 |  |  |
| **Cancer Group** |  |  |  |  |  |  |
| Colorectal | 39 (6.3) | 2 (0.3) | — |  | — |  |
| Others | 88 (14.3) | 8 (1.3) | 1.79 (0.29,10.95) | 0.53 | 1.13 (0.18,7.25) | 0.89 |
| Lymphoid/Haematopoietic | 176 (28.5) | 102 (16.5) | 11.08 (2.6,47.18) | **0.001** | 6.68 (1.21,36.91) | **0.03** |
| Non-melanoma skin | 74 (12.0) | 3 (0.5) | 0.79 (0.13,4.97) | 0.80 | 1.02 (0.13,8.09) | 0.99 |
| Breast | 10 (1.6) | 1 (0.2) | 1.95 (0.16,23.84) | 0.60 | 1.26 (0.15,10.36) | 0.83 |
| Upper GI and hepatobiliary | 61 (9.9) | 6 (1.0) | 1.92 (0.37,9.94) | 0.44 | 1.54 (0.25,9.71) | 0.64 |
| Prostate | 26 (4.2) | 5 (0.8) | 3.75 (0.67,20.97) | 0.13 | 5.26 (0.75,37.03) | 0.09 |
| Lung | 10 (1.6) | 2 (0.3) | 3.9 (0.51,29.63) | 0.19 | 2.36 (0.22,25.58) | 0.48 |
| Melanoma | 4 (0.6) | 0 (0.0) |  |  |  |  |
| **Time from the First Cancer** |  |  |  |  |  |  |
| <6m | 181 (29.3) | 78 (12.6) | — |  |  |  |
| 6–12m | 51 (8.3) | 10 (1.6) | 0.47 (0.23,0.97) | **0.04** |  |  |
| 1–2y | 38 (6.2) | 10 (1.6) | 0.64 (0.29,1.42) | 0.27 |  |  |
| 2–5y | 85 (13.8) | 19 (3.1) | 0.53 (0.27,1.05) | 0.07 |  |  |
| >5y | 133 (21.6) | 12 (1.9) | 0.23 (0.11,0.47) | **<0.001** |  |  |
| **Years since the First Cancer Diagnostic Code, per year** | 1.3 (0.2,5.6) | 0.4 (0.1,1.9) | 0.88 (0.81,0.96) | **0.004** |  |  |
| **Active Chemotherapy Days in the Last Year, per 30 days** | 0 (0, 127) | 71 (0, 177) | 1.09 (1.03,1.14) | **0.001** |  |  |
| **Active Chemotherapy in Last Year** | 240 (38.9) | 90 (14.6) | 2.52 (1.62,3.94) | **<0.001** |  |  |
| **1-year-look-back Charlson Score, per 1 score** | 2 (1,3) | 1 (1,2) | 0.84 (0.73,0.96) | **0.009** |  |  |
| **IMD Percentile, per 1 percentile (3 unknown)** | 76.8 (57.5,88.4) | 78.5 (56.1,90.4) |  |  |  |  |
| **Healthcare & Clinical History:** |  |  |  |  |  |  |
| **Number of Positive Urine Cultures with Enterococcus faecalis/faecium in Last Year, per 1 culture** | 0 (0,0) | 0 (0,0) | 0.9 (0.44,1.83) | 0.77 |  |  |
| **Positive CSU Cultures in Last Year** | 37 (6.0) | 10 (1.6) | 1.04 (0.51,2.11) | 0.92 |  |  |
| **1-year-look-back Frailty Score, per 10 scores** | 10.7 (5.2,18.0) | 9.5 (5.2,17.8) | 0.93 (0.75,1.15) | 0.48 | 1.40 (1.02,1.93) | **0.04** |
| **Days in Hospital before Collection Date in Last Year, per 10 days** | 18.5 (6.0,39.0) | 31.0 (15.0,63.0) | 1.13 (1.07,1.19) | **<0.001** |  |  |
| **Days in ICU before Collection Date in Last Year, per 1 day** | 0 (0,0) | 0 (0,1) | 1.02 (0.99,1.06) | 0.15 |  |  |
| **Blood Infection Onset** |  |  |  |  |  |  |
| Within 48h of Current Admission (Community) | 212 (34.4) | 23 (3.7) | — |  | — |  |
| Beyond 48h of Current Admission (Hospital) | 276 (44.7) | 106 (17.2) | 3.42 (2.04,5.74) | **<0.001** |  |  |
| **Antibiotics Exposure:** |  |  |  |  |  |  |
| **Days on Non-Vancomycin Antibiotics in Last Year, per 10 days** | 0 (0,9) | 0 (0,25) | 1.2 (1.1,1.3) | **<0.001** |  |  |
| **Days on Vancomycin in Last Year, per 1 day** | 0 (0,0) | 0 (0,0) | 1.13 (1.07,1.19) | **<0.001** |  |  |
| **Days Since Last Antibiotic Used in Last Year, per 30 days** | 365 (5,365) | 365(1,365) | **Non-linear** | |  |  |
| **Previous Resistance:** |  |  |  |  |  |  |
| **Previous Resistance in the Last Year** |  |  |  |  |  |  |
| No Previous AST Result/Positive Isolates, from Enterococcus faecalis/faecium in Any Cultures | 399 (64.7) | 74 (12.0) | — |  | — |  |
| Resistance to Vancomycin in Enterococcus faecalis/faecium in Any Cultures | 7 (1.1) | 28 (4.5) | 20.82 (8.89,48.78) | **<0.001** | 16.94 (6.04,47.54) | **<0.001** |
| Resistance Only to Non-Vancomycin in Enterococcus faecalis/faecium in Any Cultures³ | 72 (11.7) | 25 (4.1) | 1.87 (1.1,3.19) | **0.02** | 1.77 (1.00,3.14) | **0.05** |
| Susceptible to All Tested Antimicrobials in Enterococcus faecalis/faecium in Any Cultures | 10 (1.6) | 2 (0.3) | 1.08 (0.23,5.03) | 0.92 | 2.57 (0.50,13.13) | 0.26 |
| **Microbiological Cultures Sent:** |  |  |  |  |  |  |
| **Number of Urine Samples Taken in Last Year, per 10 cultures** | 2 (1,5) | 3 (1,6) | 1.97 (1.1,3.55) | **0.03** |  |  |
| **Number of Neutrophil Tests in Last Year, per 10 tests** | 22 (11,42) | 37 (21,62) | 1.17 (1.11,1.24) | **<0.001** |  |  |
| **Number of Blood Samples Taken in Last Year, per 1 sample Study** | 3 (1,9) | 12 (5,23) | 1.06 (1.04,1.07) | **<0.001** |  |  |
| **Timeline:** |  |  |  |  |  |  |
| **Calendar Days of Collection Date Since 01/04/2015, per year** | 2,055 (1,342,2,868) | 1,692 (699,2,396) | 0.87 (0.81,0.95) | **0.001** | 0.85 (0.77,0.93) | **<0.001** |
| ¹n (%); Median (Q1, Q3) | | | | | | |
| ²Abbreviations: CI = Confidence Interval, OR = Odds Ratio | | | | | | |
| ³Equivalent to fully susceptible to Vancomycin and resistant to any non-Vancomycin in this dataset. | | | | | | |

Table S9: Associations (odds ratios and 95% confidence intervals) between potential risk factors and *Enterococcus faecalis/faecium*-Vancomycin in univariable and multivariable backward selection models.

**References**

1.	Ferguson J, Maturo F, Yusuf S, et al. Population attributable fractions for continuously distributed exposures. Epidemiologic Methods. 2020;9(1):20190037.

2.	Klose M, Zivich PN, Cole SR. Revisiting the Population Attributable Fraction. Epidemiology. 2025;36(4):482–486.
